# High Sensitivity in Spontaneous Intracranial Hemorrhage Detection from Emergency Head CT Scans Using Meta-Learning Approach

**DOI:** 10.1101/2024.05.28.24308084

**Authors:** Juuso Takala, Heikki Peura, Riku Pirinen, Katri Väätäinen, Sergei Terjajev, Ziyuan Lin, Rahul Raj, Miikka Korja

## Abstract

Spontaneous intracranial hemorrhages have a high disease burden. Due to increasing medical imaging, new technological solutions for assisting in image interpretation are warranted. We developed a deep learning (DL) solution for spontaneous intracranial hemorrhage detection from head CT scans. The DL solution included four base convolutional neural networks (CNNs), which were trained using 300 head CT scans. A metamodel was trained on top of the four base CNNs, and simple post processing steps were applied to improve the solution’s accuracy. The solution performance was evaluated using a retrospective dataset of consecutive emergency head CTs imaged in ten different emergency rooms. 7797 head CT scans were included in the validation dataset and 118 CT scans presented with spontaneous intracranial hemorrhage. The trained metamodel together with a simple rule-based post-processing step showed 89.8% sensitivity and 89.5% specificity for hemorrhage detection at the case-level. The solution detected all 78 spontaneous hemorrhage cases imaged presumably or confirmedly within 12 hours from the symptom onset and identified five hemorrhages missed in the initial on-call reports. By using a limited amount of training data, a meta-learning approach and a simple rule-based post-processing step, clinicians can develop high-accuracy deep learning solutions for clinical imaging diagnostics.

## INTRODUCTION

Spontaneous intracerebral (ICH) and subarachnoid hemorrhage (SAH) account for up to one-third of all strokes ^1^. Both ICH and SAH are associated with high morbidity and mortality rates ^2,3^, and intraventricular hemorrhage (IVH) also commonly occurs with either condition. Due to the high disease burden of ICH and SAH, prompt and accurate diagnosis is critical, along with quickly initiated therapeutic actions ^4,5^.

Since non-contrast head CT scans (NCCTs) have high sensitivity for detecting acute intracranial blood within 12 hours from symptom onset, the diagnosis of acute intracranial hemorrhages is based on emergent NCCTs ^6,7^. The number of medical imaging studies is increasing globally ^8–10^. At the same time, there are increasing concerns regarding the fatigue of radiologists and its effect on diagnostic accuracy ^11,12^. Therefore, new technological solutions to assist clinicians and radiologists with rapid and accurate interpretation of imaging studies could alleviate this issue.

According to a recent review article ^13^, multiple algorithms have been developed for this task. However, the performance metrics of these solutions are poorly reported, and often suboptimal for clinical use.

Recently, we trained a model that identified SAH with high sensitivity, although the false positive rate was also high ^14^. To overcome this false positive problem, the number of images required for training a deep learning model can be exceptionally high. Based on these premises, we aimed to develop with limited amount of training data a combination of multiple deep learning (DL) algorithms that would detect spontaneous intracranial hemorrhages, namely ICH, SAH and IVH, with a high sensitivity and low false positive rate. Given the inherent diagnostic limitation of modern head CT scanners in identifying subacute blood (accuracy highest in early imaging) ^6,15^, we aimed to train the solution to give optimal results when applied to cases with acute hemorrhages. To evaluate the solution’s performance, we gathered a comprehensive dataset of emergency NCCT scans from different emergency rooms, imaged using various scanner devices.

## MATERIALS AND METHODS

The institutional review board of Helsinki University Hospital (HUH) approved the study and granted a waiver for acquiring informed consents (HUS/365/2017; HUS/163/2019; HUS/190/2021).

According to the Finnish legislation, no ethics committee approval is needed for retrospective studies using registry or archive data. We conducted study reporting following the Standards for Reporting of Diagnostic Accuracy Study (STARD) and Checklist for Artificial Intelligence in Medical Imaging (CLAIM) guidelines ^16,17^.

### Deep Learning Solution

Our solution included four base U-Nets ^18^ and a metamodel that was trained on top of the base U-Nets. We trained one ICH U-Net, one IVH U-Net and two SAH U-Nets to detect ICH, SAH and IVH. The training and performance metrics of one of the two SAH U-Nets have been described before ^14^. In short, this SAH model was developed with 120 NCCTs, of which 98 included SAH, and 22 were negative for SAH. We trained the new ICH, SAH and IVH U-Nets with 180 NCCTs collected from HUH picture archiving and communication systems. For training the three new U-Nets, we segmented only the bleeding type of interest present in the NCCTs. The new SAH U-Net was developed in order improve the detection of focal SAHs.

After training the base U-Net models, we trained meta-learning model with 55 NCCTs randomly sampled from the 180 previously mentioned NCCTs. For these 55 NCCTs, we segmented all three bleeding types. All NCCTs used for model training were imaged prior to October 2021, and therefore did not overlap with our validation dataset.

The training dataset of the three new U-Nets consisted of 63, 50 and 67 head NCCT MPR reformates (with 512 x 512 dimensions, 3 mm slice thickness) for ICH, IVH, and SAH, respectively. All patients were imaged and treated at HUH prior to October 2021. Segmentations were done using a Philips IntelliSpace Discovery (Philips Healthcare, 3000 Minuteman Rd, Andover, MA) and 3D Slice^19^. Eventually, the segmentation masks were saved in a binary format with each hemorrhage subtype segmentation on a unique label.

The DL solution performs semantic segmentation (i.e. predicting pixel-wise probability for the presence of intracranial hemorrhage). After the base U-Net and metamodel development, the system was implemented with a post-processing pipeline for improving the solution detection accuracy. The hemorrhage detection is based on the segmentation output and all positive segmentations exceeding the final post processing threshold are considered positive for hemorrhage.

The detailed description of DL solution training and post-processing pipeline is provided in the article supplementary material (Supplementary Materials and Methods, Supplement 1). The code for developing the solution and performing the inference tasks are available at GitHub (https://github.com/Juusotak/Intracranial_hemorrhage_metalearning).

### Validation Dataset

To simulate a real-world emergency imaging setting, we collected a retrospective dataset from 10 different hospitals in the HUH catchment area, which covers over 1,700,000 inhabitants. The validation dataset consisted of all consecutive emergency head NCCTs imaged between October 1^st^ and December 31^st^ 2021. In more detail, if an NCCT scan of adult patients (18 years or older) was performed either with emergent or immediate priority, it was included in the dataset. We also collected corresponding on-call radiology reports, patient reports of the emergency clinic visit, and ambulance reports, when available. In addition to these, we gathered information about patient demographics (age and sex). The time of the symptom onset and the etiology of hemorrhages were evaluated based on patient and ambulance reports. The on-call radiology reports were considered as ground truths. Primary spontaneous intracranial hemorrhages included non-traumatic aneurysmal and non-aneurysmal SAHs, non-traumatic deep and lobar ICHs, and non-traumatic IVHs. Secondary spontaneous intracranial hemorrhages included SAHs, ICHs or IVHs related to ischemic strokes and tumors, and were excluded from the analysis. The head NCCT scans in which the on-call radiologist had reported hemorrhage were independently annotated on a slice-level by two study authors (JT; Medical doctor with 3 years of experience in neuroimaging and deep learning research and KV; Radiologist in-training with 2.5 years of experience) for the presence of hemorrhage using 3D Slicer^19^. Senior study author (MK; Consultant cerebrovascular neurosurgeon with 20 years of experience) solved any conflicts between the annotators either by removing annotations from the slice or accepting the slice annotations.

### Statistical Analyses

The case- and slice-level metrics were calculated using NumPy (version 1.24.0) and SciPy (version 1.9.3) Python packages. These reported metrics included sensitivity, specificity, false positive rate, negative predictive, positive predictive value, accuracy, and 95% confidence intervals (CI) for these metrics. We used the same Python packages for calculating the validation dataset demographics and interquartile ranges (IQR).

## RESULTS

### Validation Dataset

The final validation dataset included 118 NCCTs in which on-call radiologists reported spontaneous intracranial hemorrhages, and 7679 NCCTs that were reported to be negative for hemorrhages by the on-call radiologist (Figure 1). The dataset included 4078 NCCTs of females (median age 74 years old, IQR 29 years) and 3719 NCCTs of males (median age 67 years old, IQR 27 years) (Table 1). The median age of all imaged patients was 71 years (IQR 29 years) (Table 1). The 7797 NCCTs were imaged using 12 different CT scanners from four vendors (Table 1).

**Figure 1.**
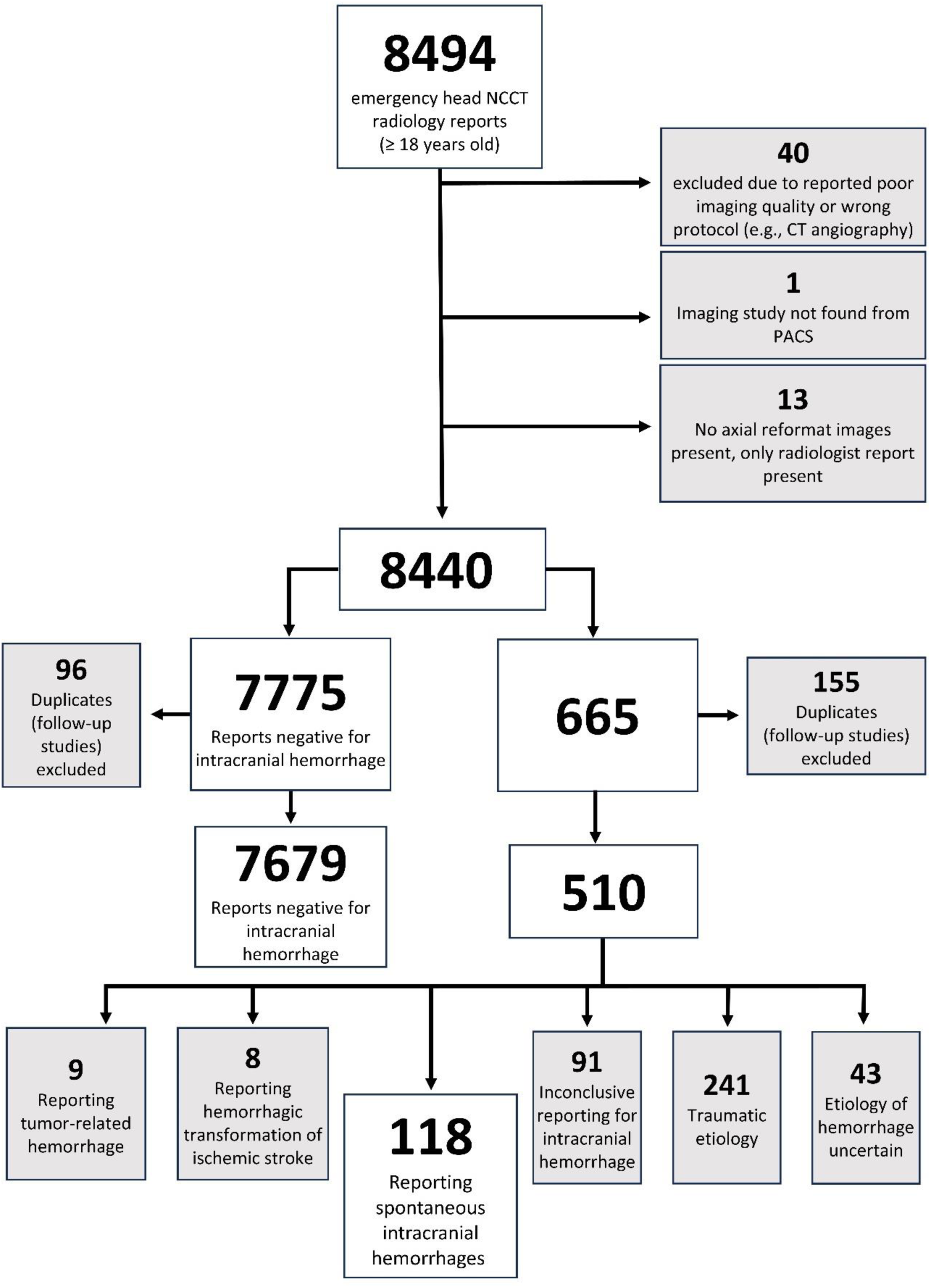
Dataset Selection. We Reviewed all 8494 on-call report for the NCCTs. The final dataset included 118 NCCTs with spontaneous hemorrhage and 7679 NCCTs which were reported negative for hemorrhage in the on-call report. The dataset did not include head NCCTs from patients who were already admitted to a hospital ward before imaging. Also, if a single patient was imaged multiple times during different emergency clinic visits during the 3 months period, all first scans (excluding follow up scans) were included in the dataset.

**Table 1.** Validation dataset patient demographics and CT scanner devices used in imaging. IQR = interquartile range.

|  | Hemorrhage | Negative | Total |
| --- | --- | --- | --- |
| **Age Median [IQR]** | 72 [20] years | 71 [29] years | 71 [29] years |
| **Female** | 57 | 4021 | 4078 |
| **Male** | 61 | 3658 | 3719 |
| **CT Scanners** |  |  |  |
| **Siemens** |  |  |  |
| Somatom Force | 25 | 587 | 612 |
| Somatom Definition Edge | 22 | 386 | 408 |
| Somatom Definition AS | 21 | 1427 | 1448 |
| Somatom Definition AS+ | 11 | 1435 | 1446 |
| Somatom Definition Flash | 6 | 679 | 685 |
| **Siemens Healthineers** |  |  |  |
| Somatom go.Top | 9 | 582 | 591 |
| Somatom X.cite | 1 | 51 | 52 |
| **GE Medical Systems** |  |  |  |
| Revolution Evo | 11 | 1147 | 1158 |
| Revolution HD | 7 | 796 | 803 |
| Discovery CT750 HD | 1 | 5 | 6 |
| LightSpeed VCT | 0 | 3 | 3 |
| **Toshiba** |  |  |  |
| Aquilon Prime | 4 | 581 | 585 |

Of the 118 NCCTs in which on-call radiologists had reported hemorrhage, 59 were imaged within 12 hours, 19 presumably within 12 hours and 17 presumably within 12 to 24 hours or confirmedly within 12 to 24 hours from the symptom onset (Table 2). Of the 59 early imaged (within 12 hours) spontaneous intracranial hemorrhages, 32 contained only one type of hemorrhage, 22 two types and 6 three types. Overall, 49 NCCTs included only one type of hemorrhage, 38 included two hemorrhage types and 10 images included three types (Table 2). The dataset also included one acute subdural hemorrhage without known head trauma.

**Table 2.** Hemorrhage Types in Validation Dataset. Counts of different bleed types and combinations of bleed types in dataset. ICH = intracerebral hemorrhage, IVH = intraventricular hemorrhage, SAH = subarachnoid hemorrhage, ASDH = acute subdural hemorrhage

|  |  |  |
| --- | --- | --- |
| **Symptom onset time confirmed < 12h** | ICH | 27 |
|  | SAH | 4 |
|  | ICH + IVH | 14 |
|  | ICH + SAH | 6 |
|  | SAH + IVH | 2 |
|  | ICH + IVH + SAH | 6 |
|  | Total | 59 |
| **Symptom onset time presumably < 12h** | ICH | 3 |
|  | IVH | 1 |
|  | ASDH | 1 |
|  | ICH + IVH | 5 |
|  | ICH + SAH | 2 |
|  | SAH + IVH | 4 |
|  | ICH + IVH + SAH | 3 |
|  | Total | 19 |
| **Symptom onset time presumably 12 - 24h**<br>**or confirmed 12- 24h** | ICH | 8 |
|  | SAH | 3 |
|  | ICH + IVH | 3 |
|  | ICH + SAH | 2 |
|  | ICH + IVH + SAH | 1 |
|  | Total | 17 |
| **Symptom onset time 24h - 7 days** | ICH | 4 |
|  | SAH | 5 |
|  | ICH + IVH | 2 |
|  | ICH + SAH | 3 |
|  | Total | 14 |
| **Symptom onset time > 7 days or unclear** | ICH | 7 |
|  | SAH | 1 |
|  | ICH + SAH | 1 |
|  | Total | 9 |

### Technical Performance

Figure 2 depicts the time range taken for analyzing all slices in one head axial 3 mm NCCT scan (median number of slices 53). The median time of analyzing all slices in a single scan was 6.7 seconds (range from 4.2 to 33.1 seconds). The analyzing procedure consisted of multiple steps, including reading the DICOM files, processing the imaging data, predicting the presence of a hemorrhage, post-processing step, and saving the predictions in a NIfTI file. Example of DL solution output is presented in Figure 3.

**Figure 2.**
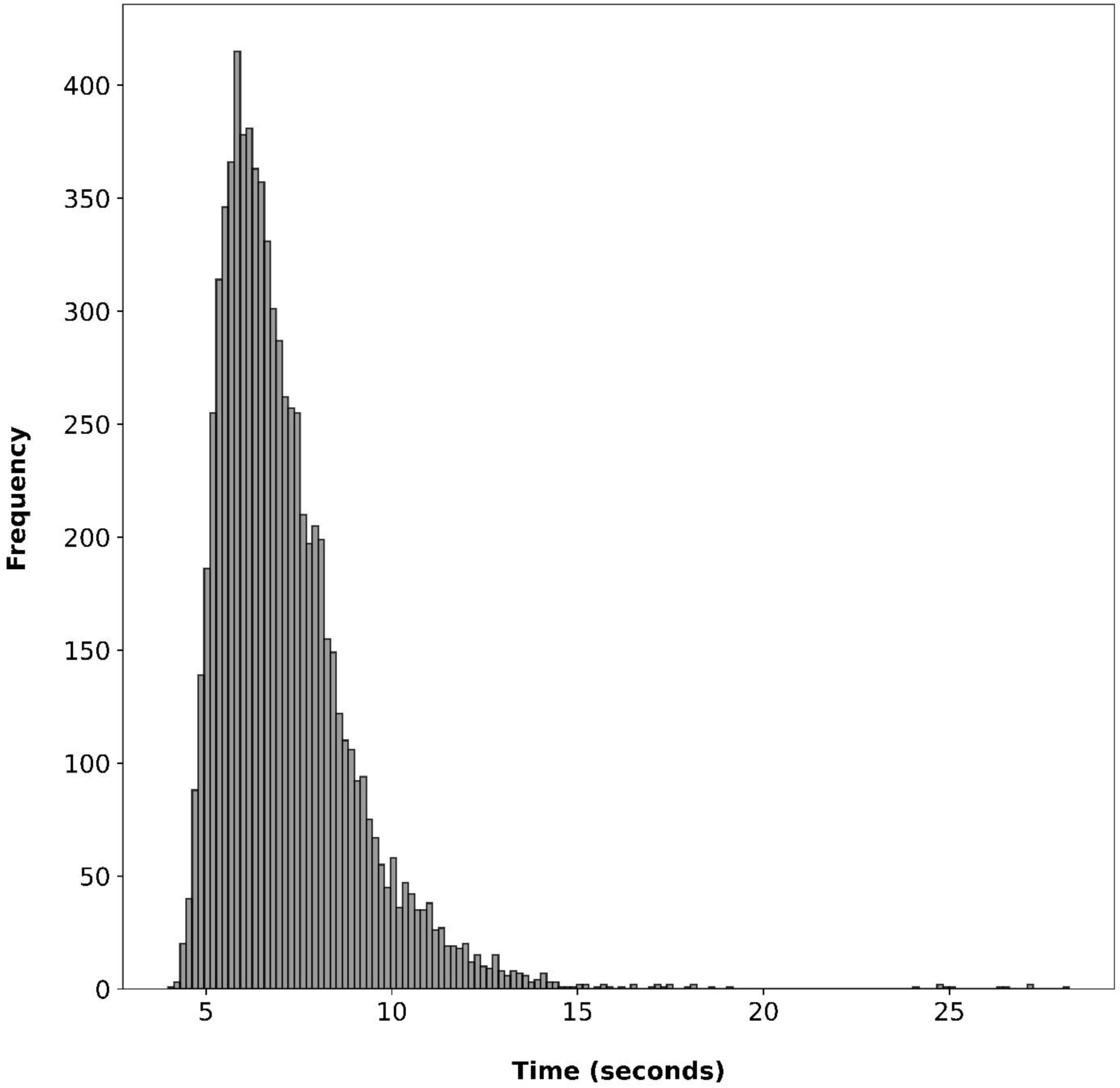
**Deep Learning Solution’s Run Times**. Histogram of inference times for scans using Nvidia Tesla V100 graphics processing unit (GPU) for running the inference task in Microsoft Azure Machine Learning Studio. The median time for analyzing a single scan was 6.7 seconds and 95 % of the scans could be analyzed within 10.6 seconds. The run time included processing of all slices in 3 mm axial reformat scan and saving the prediction.

**Figure 3.**
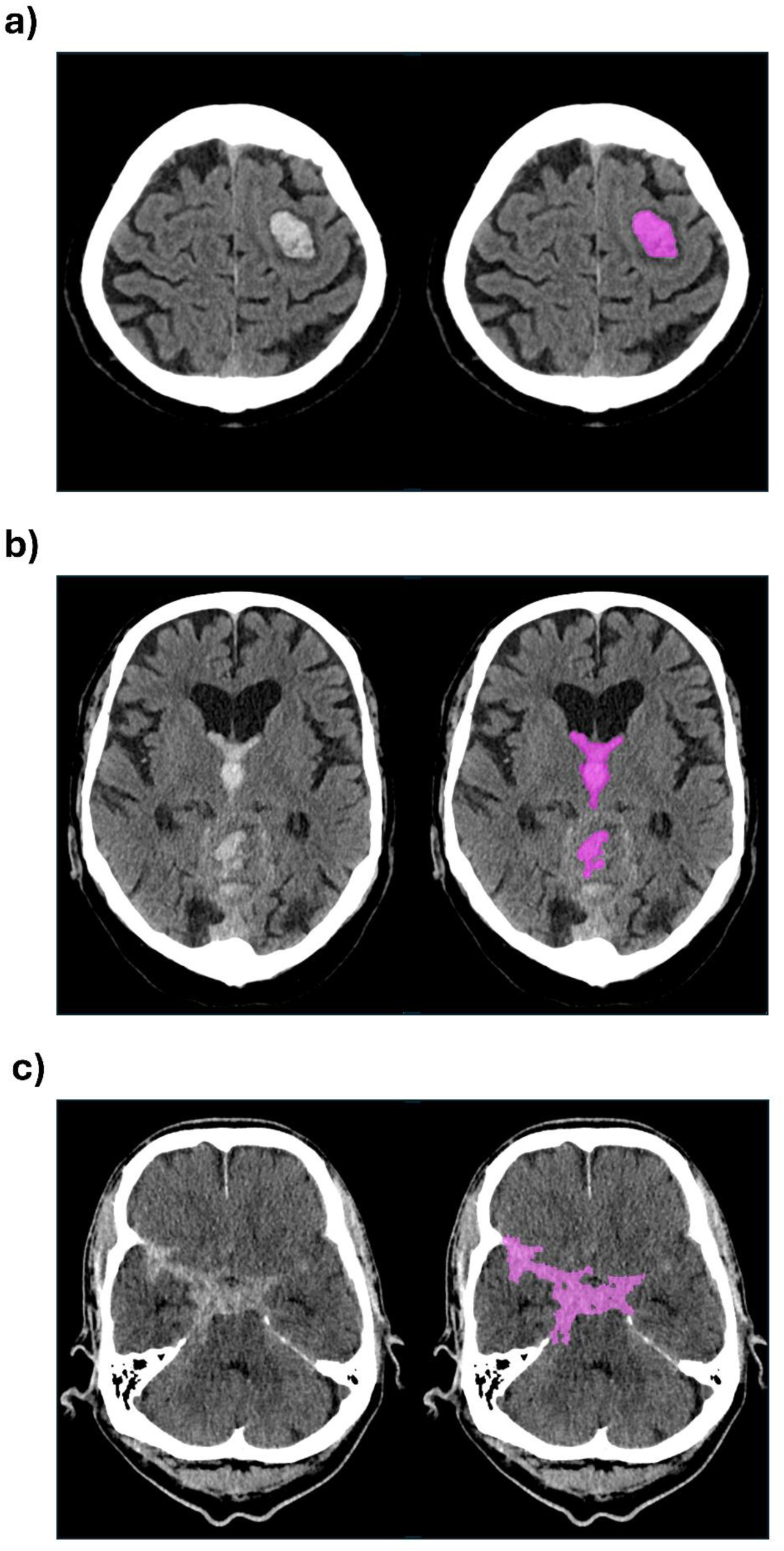
True Positive Detections of Hemorrhage and Segmentations. (a) intracerebral hemorrhage, (b) intraventricular hemorrhage and (c) subarachnoid hemorrhage in axial non-contrast head CT 3 mm reformat slice. The left sided image in each panel shows the original CT scan slice and the right sided image displays the DL solution’s detection of hemorrhage in the slice with purple color.

### Performance of Metamodel and Base U-Nets

Table 3 represents the case-level performance metrics for the four base U-Nets and the metamodel. The sensitivity and specificity of the metamodel was 92.4% (95% CI, 87.6 - 97.2%) and 53.8% (95% CI, 52.7 - 54.9%), respectively. The case-level false positive rate of the metamodel was 46.2% (95% CI, 45.1 - 47.3%). Both SAH1 and SAH2 base U-Nets had 100% sensitivity for hemorrhage detection at the case-level.

**Table 3.** Case-level performance of the base U-Nets and metamodel for all scans in validation dataset. SAH1 base U-Net is model trained during this study. SAH2 is model which development and validation was previously described by Thanellas et al ^14^. The best achieved metrics are bolded. TP = true positive, TN = true negative, FP = false positive, FN = false negative, FPR = false positive rate, NPV = negative predictive value, PPV = positive predictive value, CI = confidence interval.

|  | TP | TN | FP | FN | Sensitivity [95% CI] | Specificity [95% CI] | FPR [95% CI] | NPV [95% CI] | PPV [95% CI] | Accuracy [95% CI] |
| --- | --- | --- | --- | --- | --- | --- | --- | --- | --- | --- |
| **Metamodel - No Post-processing** | 109 | 4131 | 3548 | 9 | 92.4 % [87.6 - 97.2 %] | **53.8 % [52.7 - 54.9 %]** | **46.2 % [45.1 - 47.3 %]** | 99.8 % [99.6 - 99.9 %] | **3.0 % [2.4 - 3.5 %]** | **54.4 % [53.3 - 55.5 %]** |
| **ICH Base U-Net** | 115 | 2974 | 4705 | 3 | 97.5 % [94.6 - 100.0 %] | 38.7 % [37.6 - 39.8 %] | 61.3 % [60.2 - 62.4 %] | 99.9 % [99.8 - 100.0 %] | 2.4 % [2.0 - 2.8 %] | 39.6 % [38.5 - 40.7 %] |
| **IVH Base U-Net** | 110 | 2221 | 5458 | 8 | 93.2 % [88.7 - 97.8 %] | 28.9 % [27.9 - 29.9 %] | 71.1 % [70.1 - 72.1 %] | 99.6 % [99.4 - 99.9 %] | 2.0 % [1.6 - 2.3 %] | 29.9 % [28.9 - 30.9 %] |
| **SAH1 Base U-Net** | 118 | 1263 | 6416 | 0 | **100.0 % [100.0 - 100.0 %]** | 16.4 % [15.6 - 17.3 %] | 83.6 % [82.7 - 84.4 %] | **100.0 % [100.0 - 100.0 %]** | 1.8 % [1.5 - 2.1 %] | 17.7 % [16.9 - 18.6 %] |
| **SAH2 Base U-Net** | 118 | 922 | 6757 | 0 | **100.0 % [100.0 - 100.0 %]** | 12.0 % [11.3 - 12.7 %] | 88.0 % [87.3 - 88.7 %] | **100.0 % [100.0 - 100.0 %]** | 1.7 % [1.4 - 2.0 %] | 13.3 % [12.6 - 14.1 %] |

Supplementary Table 1 (Supplement 1) presents slice-level performance metrics for the base U-Nets and metamodel. The sensitivity and specificity of the metamodel was 71.7% (95% CI, 69.7 - 73.7%) and 98.2% (95% CI, 98.1 - 98.2%), respectively. The slice-level false positive rate of the metamodel was 1.8% (95% CI, 1.8 - 1.9%). At the slice-level, the ICH, IVH, SAH1 and SAH2 base U-Nets had false positive rates of 4.3% (95% CI, 4.3 - 4.4%), 4.3 % (95% CI, 4.3 - 4.4%), 9.9% (95% CI, 9.8 - 10.0%) and 10.1% (95% CI, 10.0 - 10.2%), respectively.

### Metamodel Performance with Full Post-processing

Table 4 represents the case-level performance metrics of the metamodel with full post-processing. The case-level sensitivity and specificity of the metamodel were 89.8% (95% CI, 84.4 - 95.3%) and 89.5% (95% CI, 88.8 - 90.2%), respectively. The case-level false positive rate for the metamodel was 10.5% (95% CI, 9.8 - 11.2%).

**Table 4.** Case-level performance metrics of the metamodel with full post-processing. The performance metrics for scans with hemorrhage are also stratified according to delay from symptom onset to imaging. TP = true positive, TN = true negative, FP = false positive, FN = false negative, FPR = false positive rate, NPV = negative predictive value, PPV = positive predictive value, N/A = not applicable, CI = confidence interval.

|  | TP | TN | FP | FN | Sensitivity % [95% CI] | Specificity % [95% CI] | FPR % [95% CI] | NPV % [95% CI] | PPV % [95% CI] | Accuracy % [95% CI] |
| --- | --- | --- | --- | --- | --- | --- | --- | --- | --- | --- |
| All Scans | 106 | 6873 | 806 | 12 | 89.8 % [84.4 - 95.3 %] | 89.5 % [88.8 - 90.2 %] | 10.5 % [9.8 - 11.2 %] | 99.8 % [99.7 - 99.9 %] | 11.6 % [9.5 - 13.7 %] | 89.5 % [88.8 - 90.2 %] |
| Symptom onset time confirmed < 12h | 59 |  |  | 0 | 100 % [100 - 100 %] | N/A | N/A | N/A | N/A | 100 % [100 - 100 %] |
| Symptom onset time presumably < 12h | 19 |  |  | 0 | 100 % [100 - 100 %] | N/A | N/A | N/A | N/A | 100 % [100 - 100 %] |
| Symptom onset time presumably 12 – 24h<br>or Confirmed 12 - 24h | 13 |  |  | 4 | 76.5 % [56.3 - 96.6 %] | N/A | N/A | N/A | N/A | 76.5 % [56.3 - 96.6 %] |
| Symptom onset time 24h - 7 days | 10 |  |  | 4 | 71.4 % [47.8 - 95.1 %] | N/A | N/A | N/A | N/A | 71.4 % [47.8 - 95.1 %] |
| Symptom onset time > 7 days or unclear | 5 |  |  | 4 | 55.6 % [23.1 - 88.0 %] | N/A | N/A | N/A | N/A | 55.6 % [23.1 - 88.0 %] |
| Negative scans |  | 6873 | 806 |  | N/A | 89.5 % [88.8 - 90.2 %] | 10.5 % [9.8 - 11.2 %] | N/A | N/A | 89.5 % [88.8 - 90.2 %] |

The metamodel showed 69.3% (95% CI, 67.3 - 71.3%) slice-level sensitivity and 99.6% (95% CI, 99.6 - 99.6%) slice-level specificity. The slice-level false positive rate of the metamodel was 0.4%

(95% CI, 0.4 - 0.4%). In other words, of the 7679 NCCTs reported negative for hemorrhage by the on-call radiologists, the solution predicted falsely positive 1594 slices out of the 408 426 slices. The slice-level performance of the metamodel with full post-processing is presented in Supplementary Table 2 (Supplement 1). Most of the false positive pixel clusters pointed out blood in normal and highly vascularized anatomical structures, e.g. sagittal sinus, cerebellar tentorium, straight sinus, and falx cerebri (Supplementary Figures 1 and 2, Supplement 1). Detailed summaries of case- and slice-level performance metrics for each component of the DL solution are provided in Supplementary Figures 3 and 4 (Supplement 1).

### Metamodel Performance by Time of Symptom Onset

The metamodel with full post-processing detected hemorrhages in 59 out of 59 patients (sensitivity 100.0%) who were imaged within 12 hours from symptom onset (Table 4). For 19 patients imaged most likely (not 100% certain) within 12 hours from symptom onset, the solution’s sensitivity was also 100% (Table 4). An additional 18 patients were imaged 12 to 24 hours from the symptom onset. The metamodel sensitivity with full post-processing among this group was 77.8% (95 CI, 56.3 - 96.6%) (Table 4). The four missed cases included two small ICHs and two focal small SAHs (Supplementary Figures 5-8, Supplement 1). For patients imaged between 24 hours to 7 days, the sensitivity of the metamodel with full post-processing was 71.4% (95% CI, 47.8 - 95.1%). The missed hemorrhage cases included one ICH and three focal SAHs (Supplementary Figures 9-12, Supplement 1). The sensitivity for hemorrhages that were imaged after 7 days from symptom onset or had unclear symptom onset time was 55.6% (95% CI, 23.1 - 88.0%). The missed hemorrhage cases included three ICHs and one focal SAH (Supplementary Figures 13-16, Supplement 1). The sensitivity of the metamodel with full post-processing for scans in different symptom-onset time points is represented in Figure 4.

**Figure 4.**
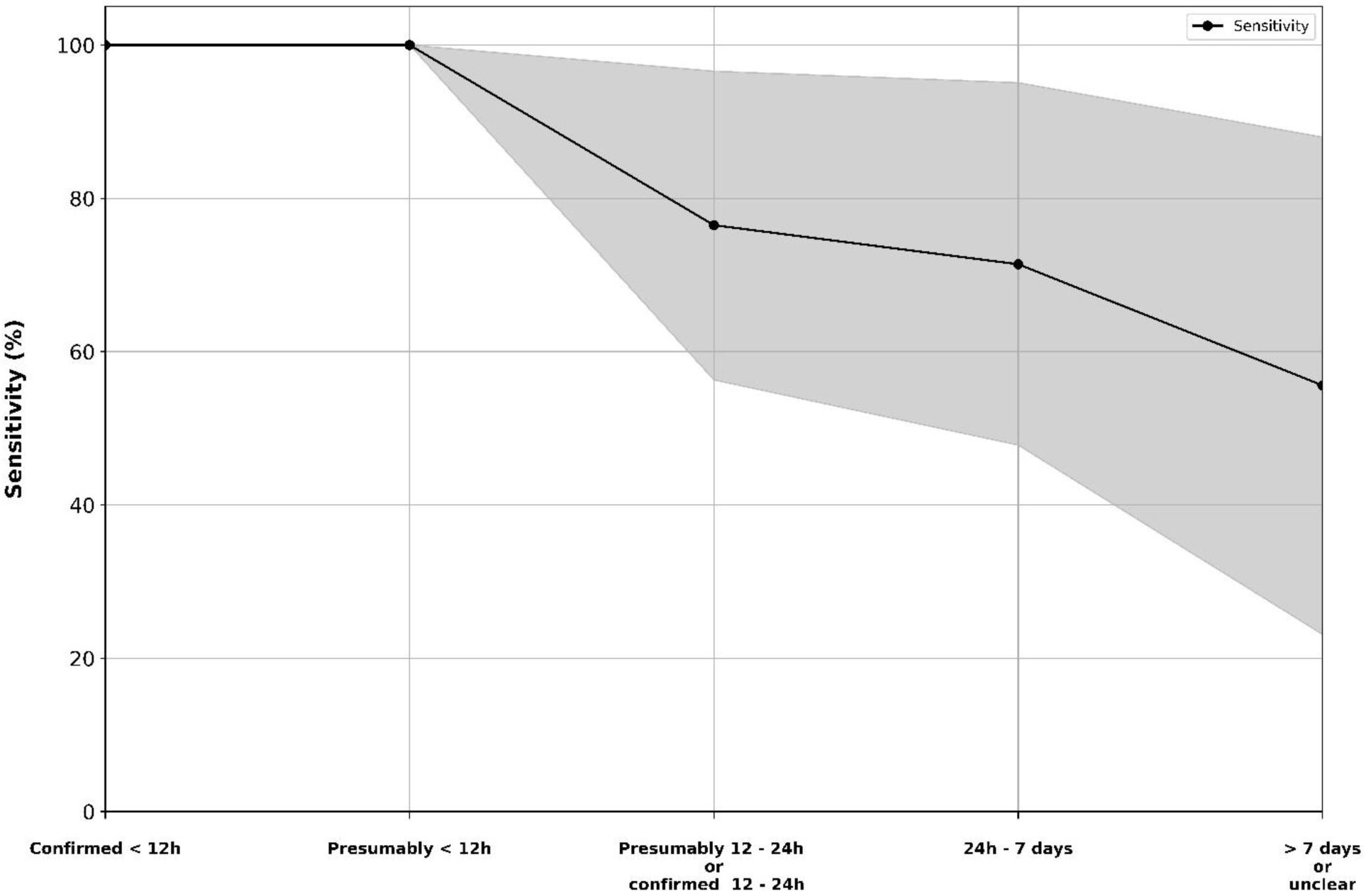
Deep Learning Solution’s Case-Level Sensitivity with Full Post-processing. Sensitivity of the deep learning solution for detecting hemorrhages at different time point groups. The shaded area in the plot represents the 95 % confidence interval.

### Cases Missed in On-Call Reports but Identified by Solution

The metamodel with full post-processing identified five spontaneous intracranial hemorrhages that were not reported in the initial on-call reports. Of these five cases, one was ICH, two were SAHs, and one was IVH. All identified hemorrhages are presented in Supplementary Figures 17-21 (Supplement 1).

## DISCUSSION

The developed DL solution detected spontaneous intracranial hemorrhages on head CT scans with 100% sensitivity up to 12 hours from symptom onset and with a low processing time per an individual NCCT scan. With the full post-processing, the solution alarmed falsely about an intracranial hemorrhage in approximately every 10^th^ true negative head NCCTs. Most of these false positive findings were present only in a few slices and in the same anatomical locations. Therefore, these small false positive pixel clusters could be relatively easily assessed as false positive findings by on-call radiologists or clinicians. Structures containing high vascularity or low pressure (slow flowing) blood, such as the choroid plexuses and intracranial sinuses, makes achieving a zero false positive rate a challenging task if a high sensitivity is a priority.

One of the DL solution’s base models has already been validated previously in both internal and external settings ^14^. Despite the model had high sensitivity for detecting SAH, there was a relatively high number of false positive findings. Due to the false positive rate, model ‘s clinical usability on its own was considered suboptimal. Therefore, we developed this new solution that combines the outputs of high-sensitivity but low-specificity base U-Net models using a meta-learning approach and post-processing steps. Whereas the lowest case-level base U-Net false positive rate was 61.3 %, the false positive case-level rate was reduced to 10.5% by applying the trained metamodel and rule-based post-processing steps. This translates to an improvement of over 80% at the case-level. The training material used in this study included only 300 head NCCTs. The development of deep learning models commonly relies on large amounts of training data, leading to a large amount of work and costs related to data preprocessing and model training. Thus, the study also suggests that meta-learning approach and development of rule-based post-processing pipeline can be used for training and optimizing clinically potential deep learning systems with limited amounts of training data.

Our study has limitations. First, the final number of true hemorrhage cases in the validation dataset can be considered low. However, the dataset represents a true consecutive patient cohort imaged during a 3-month long period in 10 different hospitals with a catchment area of around 1.7 million inhabitants. Second, the validation dataset was collected from 10 different hospitals, but the validation setting was still internal. Therefore, the solution’s results cannot be generalized outside the study country. Third, we did not have a possibility to assess our solution’s usefulness in the clinical workflow, as the solution is not an officially approved medical device. Fourth, using on-call radiologist reports as a ground truth might be considered a shortcoming, as it is recognized that some degree of error is inherent in the interpretation of medical imaging studies ^20,21^. However, since our aim was to assess if the model could improve real-world diagnostics, using the reports as ground truth was necessary. By using this approach, we were able to evaluate whether the system could identify hemorrhage cases that have been missed in a real-life situation. Fifth, since the sensitivity diminishes by the delay from symptom onset to imaging, the model suits best for acute (12 hours) cases. This may be considered as a shortcoming, even though the model was designed for emergency use and for acute cases, only.

The sensitivity of acute intracranial hemorrhage detection of our solution is essentially similar to the reported performance metrics of commercially available and clinically used solutions ^22–25^.

However, reliable comparisons between different solutions are difficult to conduct due to the lack of standardized comparison protocols and datasets. Although emergency head NCCTs interpreted by radiologists have a high sensitivity for acute blood ^6,7^, misidentification of acute intracranial hemorrhage still occurs in clinical settings. Our solution successfully detected five cases of acute spontaneous intracranial hemorrhages that were not reported in on-call reports. Due to relatively low computational costs of the 2-dimensional U-Net architecture, the solution could also be run in a local set up without requiring costly high-end graphics processing units or central processing units.

In conclusion, the developed DL solution with the post-processing pipeline had high sensitivity of detecting spontaneous intracranial hemorrhages in the acute period. This kind of solution could help to rule out spontaneous intracranial hemorrhages in a clinical setting. Even though the solution is not an officially approved medical device, it could already be used for quality assessments and research purposes.

## DATA AVAILABILITY

Finnish healthcare data for secondary use can be obtained through FINDATA (Social and Health Data Permit Authority according to the Secondary Data Act). The used healthcare data cannot be shared openly. The code for training the deep learning models and running the inference tasks are available at: https://github.com/Juusotak/Intracranial_hemorrhage_metalearning

## ACKNOWLEDGEMENTS

This work is part of the AI Head Analysis project of the CleverHealth Network ecosystem (https://www.cleverhealth.fi/en/home), and we thank the ecosystem partners for supporting the project. JT received a research grant from Maire Taposen foundation. The study project was supported by a grant from the State Research Funds (Helsinki University Hospital).

## AUTHOR CONTRIBUTIONS STATEMENT

J.T., H.P. and Z.L. developed the deep learning solution (including base model and meta-model training, and development of the post-processing pipeline). H.P., S.T., R.R. and M.K. contributed to the acquisition and curation of the training data. J.T., R.P., K.V. and M.K. contributed to the acquisition and curation of the validation dataset. J.T. and M.K. analyzed the results and wrote the main manuscript text. J.T. prepared all figures and tables. M.K. and R.R. contributed to the administration of the research project. All authors reviewed and edited the manuscript as needed and approved its final version.

