## Supplement 1 for "High Sensitivity in Spontaneous Intracranial Hemorrhage Detection from Emergency Head CT Scans Using Meta-Learning Approach"

**Supplementary Materials and Methods**

Training Data

Training dataset of the three new U-Nets consisted of 63, 50 and 67 head non-contrast head CT scan (NCCT) MPR-reformates (with 512 x 512 dimensions, 3 mm slice thickness) for intracerebral hemorrhage (ICH), intraventricular hemorrhage (IVH), and subarachnoid hemorrhage (SAH), respectively. All patients were imaged and treated at Helsinki University Hospital prior to October 2021. Segmentations were done using Philips IntelliSpace Discovery (Philips Healthcare, 3000 Minuteman Rd, Andover, MA) and 3D Slicer^1^ (http://www.slicer.org). Image data and segmentation files were saved and used in NIfTI file format. We used Hounsfield unit (HU) threshold-based method to decrease human error and to increase reproducibility. We determined an acute blood threshold value for each bleeding type; 60-90 HUs for ICH and SAH and 50-90 HUs for IVH. We only segmented the bleeding type of interest for the base model training. The segmentations for training dataset were carried out by one of the study authors (HP; Medical doctor with 3 years of experience in deep learning and neuroimaging research). To make a final agreement of the segmentation mask, all segmented NCCTs were reviewed by a neurosurgeon and/or radiologist (MK; Consultant cerebrovascular neurosurgeon with 20 years of experience, RR; Neurosurgeon in-training with 8 years of experience, ST; Consultant radiologist with 8 years of experience). After the review, segmentation masks were saved in a binary format, 0 meaning “no blood” and 1 meaning “blood”. We randomly chose 55 NCCTs scans from the base model training dataset for the meta-model training dataset. For these 55 NCCTs we segmented all three bleeding types using the same HU thresholds mentioned above. The review process after segmentation remained the same. Eventually, the segmentation masks were saved also in a binary format with each hemorrhage subtype segmentation on unique label. The final segmentation files for the metamodel training included three channels (one for each bleeding type).

Model Architectures and Training

The trained three U-Nets were two-dimensional (2D) and had five convolution blocks with residual connections both in the encoder and decoder parts. Dropout and batch normalization layers were also used. Down sampling was done using a 2D convolutional layer with a kernel size of (2,2) and stride of (2,2). Architecture for the new base models trained for detecting ICH, IVH and SAH is presented in the following figure:

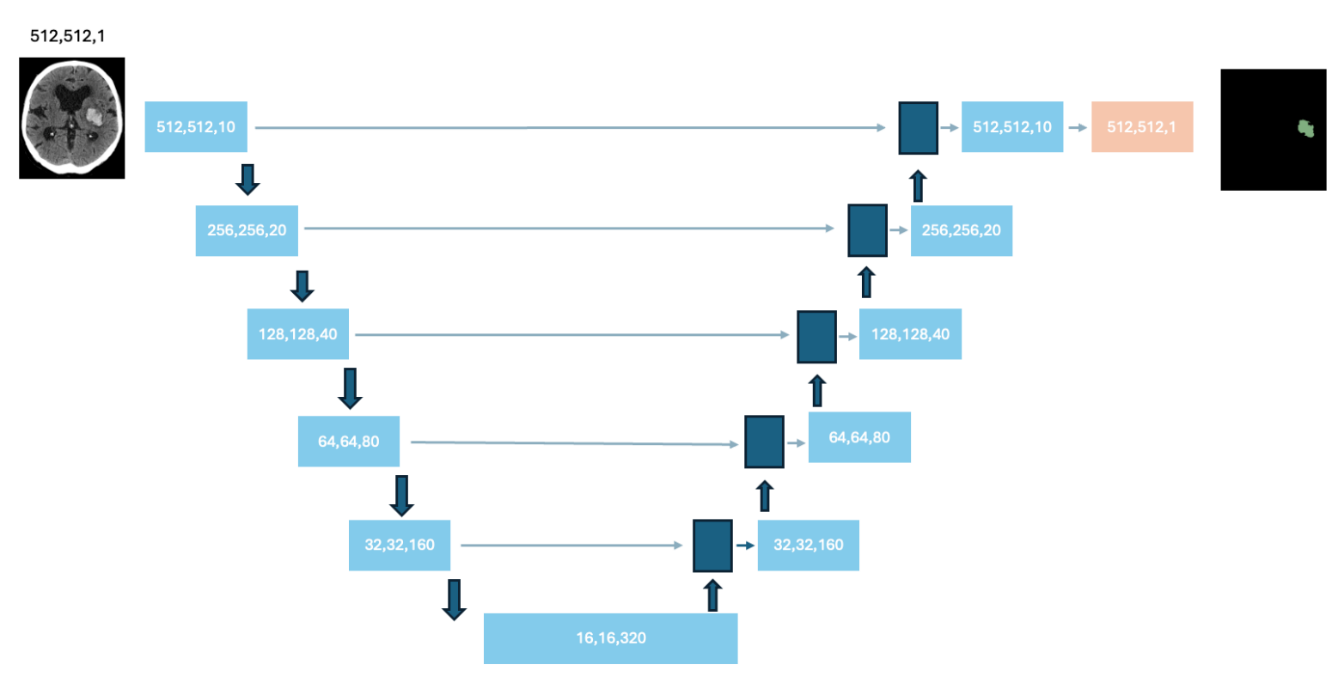

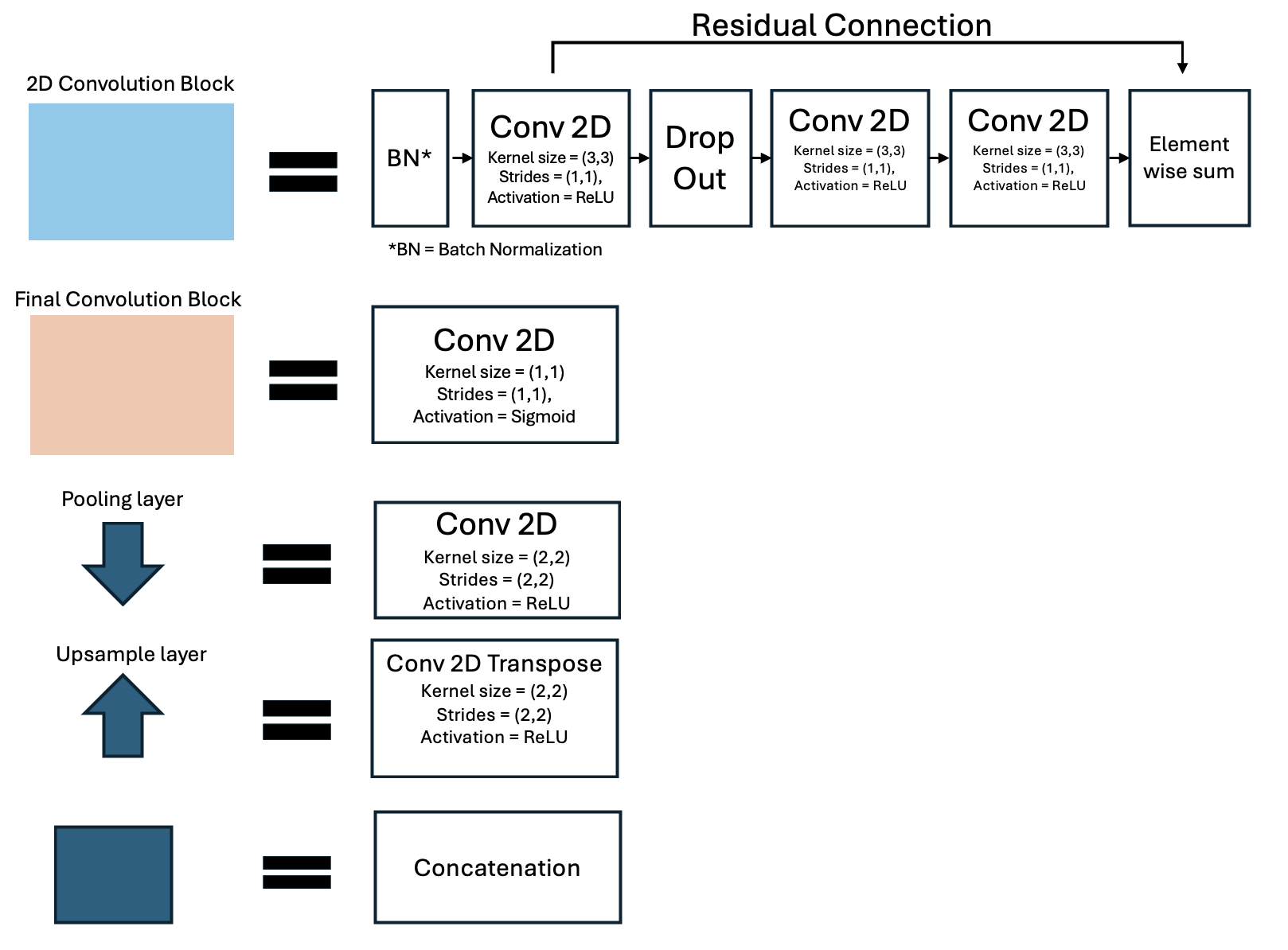

Dice loss was used as training loss for ICH and SAH U-Nets. For the IVH U-Net, we used a custom loss function where focal loss and dice loss were combined. We used random parameter initialization at the start of the training with TensorFlow’s default option (glorot_uniform).

Image data for ICH and SAH U-Nets were first clipped between -500 and 500 HUs and then normalized between 0 and 1. Image data for IVH U-Net was clipped between 0 and 200 HUs and then normalized between 0 and 1. Image data and segmentation masks were rotated 90, 180 and 270 degrees as an augmentation method during the training. Before training, each of the datasets were randomly split into training and evaluation set using 80:20 split ratio with 20% of the data reserved for evaluation purposes during training process. Training was done in Microsoft Azure using NVIDIA Tesla V100 graphics processing unit (GPU) with a batch size of 64 and dropout rate of 0.2. We used an Adam optimizer with a learning rate of 0.001. The training was set to last 200 epochs and the U-Net with the lowest evaluation loss value was selected for the final use.

We trained the metamodel after training the base U-Nets. The metamodel was trained to make the final detection and localization of the lesion suspected as hemorrhage. In other words, the metamodel was responsible for combining the multiple outputs from the separate base U-Nets for single output (i.e. detection and localization of the hemorrhage in the slice). The metamodel included a batch normalization layer and two 2D convolutional layers. A full presentation of the metamodel architecture is presented in the following figure:

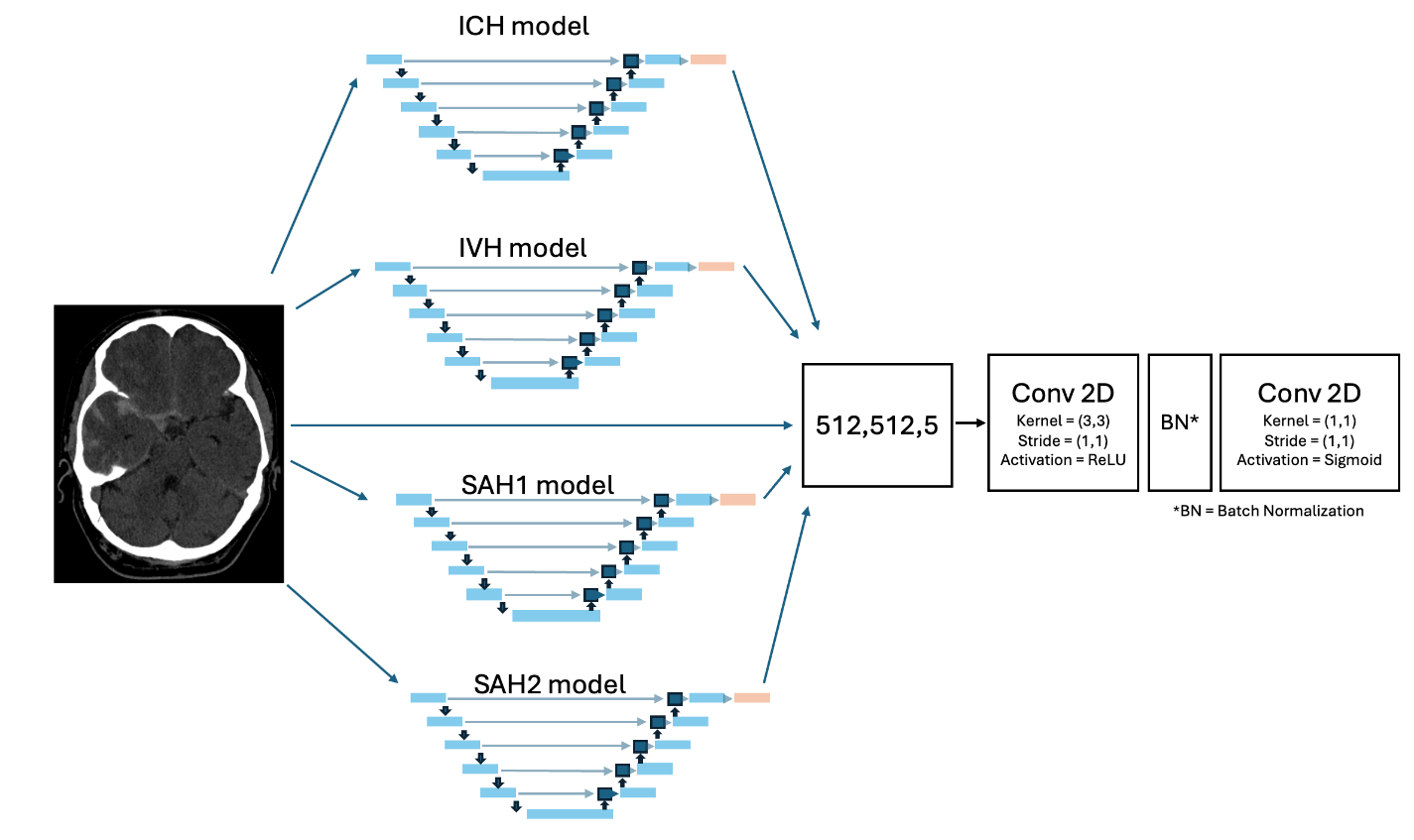

The parameters were initialized randomly at the start of the training with TensorFlow’s default option (glorot_uniform). The input of the metamodel is a 2D image with 5 channels (original CT slice and predictions from the four base U-Nets). The image channel was clipped between –500 and 500 HUs and normalized between 0 and 1.

Throughout the metamodel training phase, the weights of the base models remained unchanged (i.e. the base model weights were frozen before the metamodel training). During the training, image data and corresponding segmentation masks were rotated 90, 180 and 270 degrees as an augmentation method. Similar 80:20 split ratio was used for the metamodel training dataset. Training was done in Microsoft Azure using V100 GPU with batch size of 64, Learning rate of 0.003 and Adam optimizer. The training was set to last 100 epochs. We used Dice loss as a loss function, and the model with the lowest evaluation loss was selected for the final use. The model outputs semantic segmentation, i.e. pixel-wise probability of the presence of a hemorrhage in a single NCCT slice.

The output from the metamodel is an array with shape of 512,512,3 in which 3 channels represent the pixel-wise probability of ICH, IVH and SAH on the CT scan slice. In the inference task, pixel-wise maximum probability was selected from the metamodel output and thus the array was transformed into shape of 512,512,1.

For the inference pipeline, the combined U-Net solution included a reshaping of image data if the pixel data shape differed from 512 x 512. After resizing the image (if needed), the data was first sent to the base U-Nets with the corresponding normalizations. The base U-Net predictions were saved for later post processing steps. The predictions from the base U-Nets and the original imaging data (with clipping between -500 and 500 HU and normalization) were then directed to the metamodel for the final prediction. We used TensorFlow 2.2.0 to build and train the U-Nets and the metamodel. We used 0.5 as a cut-off value for positive pixel-wise prediction.

Adaptive Post-Processing Steps

To reduce the number of falsely predicted positive findings, we implemented post processing steps. First, we included only predictions in which segmented pixels (i.e. predicted blood) formed a cluster of a minimum 10 pixels in a single slice. A voting support from the base U-Nets was used to direct the slice further for a test time augmentation (TTA) step, or to push the slice forward without TTA. In the voting phase, all predictions from the base U-Nets were summed and compared against the metamodel predictions. If there was an area of prediction with overlap between the metamodel prediction and majority of the base U-Net predictions, the slice was classified as a strong positive. In the abovementioned comparison, the predictions from the base U-Nets were summed together pixel-wise (giving possible maximum value of 4.0). Next the summed predictions from the base U-Nets were then divided by 3.0 and finally the comparison was determined “strong positive” if there was region of overlap with the metamodel prediction and summed prediction of 1.0 or more.

TTA was applied as a post processing step only if the slice was positive, but the prediction was not classified as a strong positive, as described above. By default, the solution made the predictions for input image data without any augmentations. In the TTA step, the image data was flipped horizontally, vertically and with both axes. The augmented images were then analyzed, and the predictions were then spatially reversed to match the predictions of the original data. All predictions were then summed and divided by 4. A removal of segmented pixel clusters smaller than 10 pixels was also done after the TTA step. In the final step of the post processing, the positive prediction clusters were combined with the predictions of the base U-Nets, and if the combined cluster size exceeded 125 pixels, the segmentation was classified as a positive prediction. Of note, one 512 x 512 head NCCT slice contains 262 144 pixels.

**Supplementary Tables**

|  | Slices | TP | TN | FP | FN | Sensitivity % [95% CI] | Specificity % [95% CI] | FPR % [95% CI] | NPV % [95% CI] | PPV % [95% CI] | Accuracy % [95% CI] |
| --- | --- | --- | --- | --- | --- | --- | --- | --- | --- | --- | --- |
| Metamodel - No Post-Processing | 416222 | 1443 | 406586 | 7623 | 570 | 71.7 % [69.7 - 73.7 %] | **98.2 % [98.1 - 98.2 %]** | **1.8 % [1.8 - 1.9 %]** | 99.9 % [99.8 - 99.9 %] | **15.9 % [15.2 - 16.7 %]** | **98.0 % [98.0 - 98.1 %]** |
| ICH Base U-Net | 416222 | 1460 | 396231 | 17978 | 553 | 72.5 % [70.6 - 74.5 %] | 95.7 % [95.6 - 95.7 %] | 4.3 % [4.3 - 4.4 %] | 99.9 % [99.8 - 99.9 %] | 7.5 % [7.1 - 7.9 %] | 95.5 % [95.5 - 95.6 %] |
| IVH Base U-Net | 416222 | 693 | 396346 | 17863 | 1320 | 34.4 % [32.4 - 36.5 %] | 95.7 % [95.6 - 95.7 %] | 4.3 % [4.3 - 4.4 %] | 99.7 % [99.7 - 99.7 %] | 3.7 % [3.5 - 4.0 %] | 95.4 % [95.3 - 95.5 %] |
| SAH1 Base U-Net | 416222 | 1582 | 373337 | 40872 | 431 | 78.6 % [76.8 - 80.4 %] | 90.1 % [90.0 - 90.2 %] | 9.9 % [9.8 - 10.0 %] | 99.9 % [99.9 - 99.9 %] | 3.7 % [3.5 - 3.9 %] | 90.1 % [90.0 - 90.2 %] |
| SAH2 Base U-Net | 416222 | 1596 | 372346 | 41863 | 417 | **79.3 % [77.5 - 81.1 %]** | 89.9 % [89.8 - 90.0 %] | 10.1 % [10.0 - 10.2 %] | **99.9 % [99.9 - 99.9 %]** | 3.7 % [3.5 - 3.8 %] | 89.8 % [89.8 - 89.9 %] |

**Supplementary Table 1.** Slice-level performance of base U-Nets and metamodel for all scans in validation dataset. SAH1 base U-Net is model trained during this study. SAH2 is model which development and validation was previously described by Thanellas et al.^2^ The best achieved metrics are bolded. TP = true positive, TN = true negative, FP = false positive, FN = false negative, FPR = false positive rate, NPV = negative predictive value, PPV = positive predictive value, CI = confidence interval

|  | **Slices** | **TP** | **TN** | **FP** | **FN** | **Sensitivity % [95% CI]** | **Specificity % [95% CI]** | **FPR % [95% CI]** | **NPV % [95% CI]** | **PPV % [95% CI]** | **Accuracy % [95% CI]** |
| --- | --- | --- | --- | --- | --- | --- | --- | --- | --- | --- | --- |
| **Symptom onset time confirmed < 12h** | 3068 | 788 | 1961 | 38 | 281 | 73.7 % [71.1 - 76.4 %] | 98.1 % [97.5 - 98.7 %] | 1.9 % [1.3 - 2.5 %] | 87.5 % [86.1 - 88.8 %] | 95.4 % [94.0 - 96.8 %] | 89.6 % [88.5 - 90.7 %] |
| **Symptom onset time presumably < 12h** | 1053 | 336 | 571 | 11 | 135 | 71.3 % [67.3 - 75.4 %] | 98.1 % [97.0 - 99.2 %] | 1.9 % [0.8 - 3.0 %] | 80.9 % [78.0 - 83.8 %] | 96.8 % [95.0 - 98.7 %] | 86.1 % [84.0 - 88.2 %] |
| **Symptom onset time presumably 12 - 24 h or confirmed 12 - 24h** | 872 | 134 | 673 | 1 | 64 | 67.7 % [61.2 - 74.2 %] | 99.9 % [99.6 - 100.0 %] | 0.1 % [0.0 - 0.4 %] | 91.3 % [89.3 - 93.3 %] | 99.3 % [97.8 - 100.0 %] | 92.5 % [90.8 - 94.3 %] |
| **Symptom onset time 24h - 7 days** | 739 | 98 | 549 | 10 | 82 | 54.4 % [47.2 - 61.7 %] | 98.2 % [97.1 - 99.3 %] | 1.8 % [0.7 - 2.9 %] | 87.0 % [84.4 - 89.6 %] | 90.7 % [85.3 - 96.2 %] | 87.6 % [85.2 - 89.9 %] |
| **Symptom onset time > 7 days or unclear** | 470 | 39 | 375 | 0 | 56 | 41.1 % [31.2 - 50.9 %] | 100.0 % [100.0 - 100.0 %] | 0.0 % [0.0 - 0.0 %] | 87.0 % [83.8 - 90.2 %] | 100.0 % [100.0 - 100.0 %] | 88.1 % [85.2 - 91.0 %] |
| **False positive cases** | 43752 | 0 | 42158 | 1594 | 0 | N/A | 96.4 % [96.2 - 96.5 %] | 3.6 % [3.5 - 3.8 %] | N/A | N/A | 96.4 % [96.2 - 96.5 %] |
| **True negative cases** | 366268 | 0 | 366268 | 0 | 0 | N/A | 100.0 % [100.0 - 100.0 %] | 0.0 % [0.0 - 0.0 %] | N/A | N/A | 100.0 % [100.0 - 100.0 %] |
| **All** | 416222 | 1395 | 412555 | 1654 | 618 | 69.3 % [67.3 - 71.3 %] | 99.6 % [99.6 - 99.6 %] | 0.4 % [0.4 - 0.4 %] | 99.9 % [99.8 - 99.9 %] | 45.8 % [44.0 - 47.5 %] | 99.5 % [99.4 - 99.5 %] |

**Supplementary Table 2.** Slice-level performance metrics of the metamodel with full post-processing. The performance metrics for scans with hemorrhage are also stratified according to delay from symptom onset to imaging. TP = true positive, TN = true negative, FP = false positive, FN = false negative, FPR = false positive rate, NPV = negative predictive value, PPV = positive predictive value, N/A = not applicable, CI = confidence interval.

**Supplementary Figures**

**True Positive Scans**

**False Positive Scans**

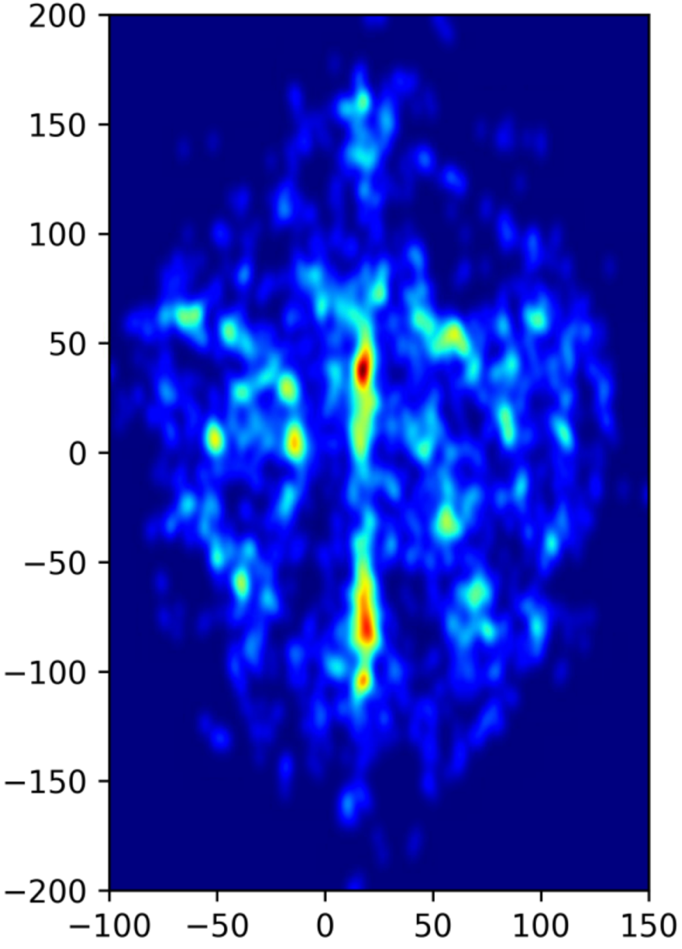

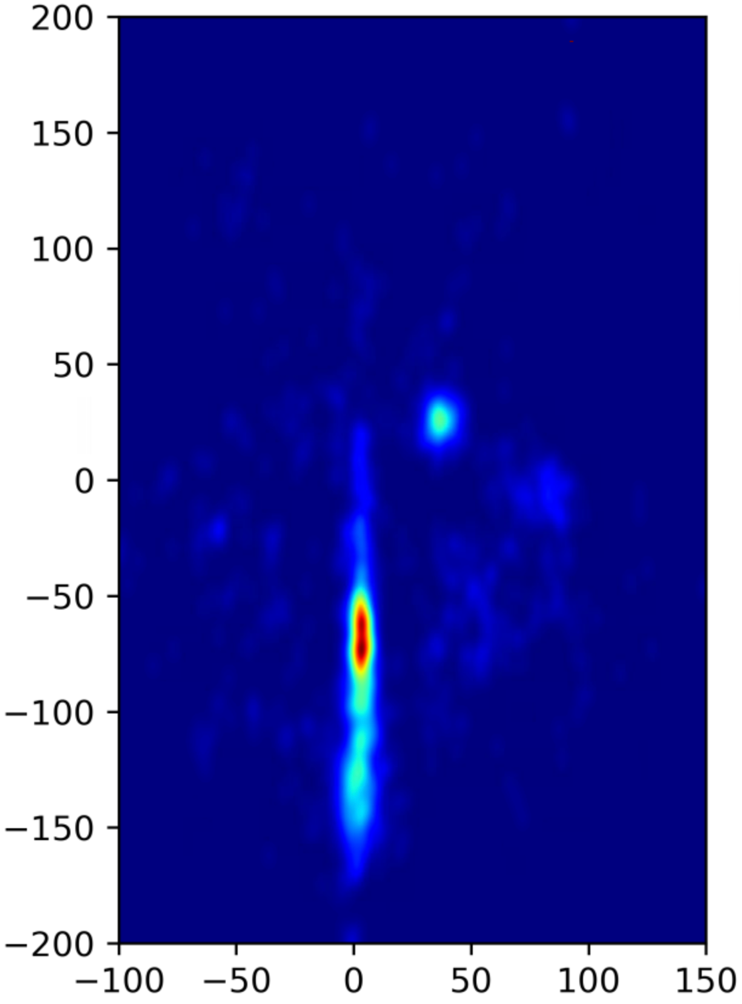

**Supplementary Figure 1. Heatmap of False Positive and True Positive Segmentations.** The heatmap presents spatial locations of the deep learning solution’s segmentations in false positive and true positive scan slices. The X- and Y- axis coordinates are calculated respective to the midpoint of the largest component in the image slice present with Hounsfield units > 150 (the cranium). The heatmap shows that the most common location for false positive segmentation is the posterior midline (e.g., falx, sagittal and straight sinus). The hemorrhage detections in true positive scans are scattered across a wider area.

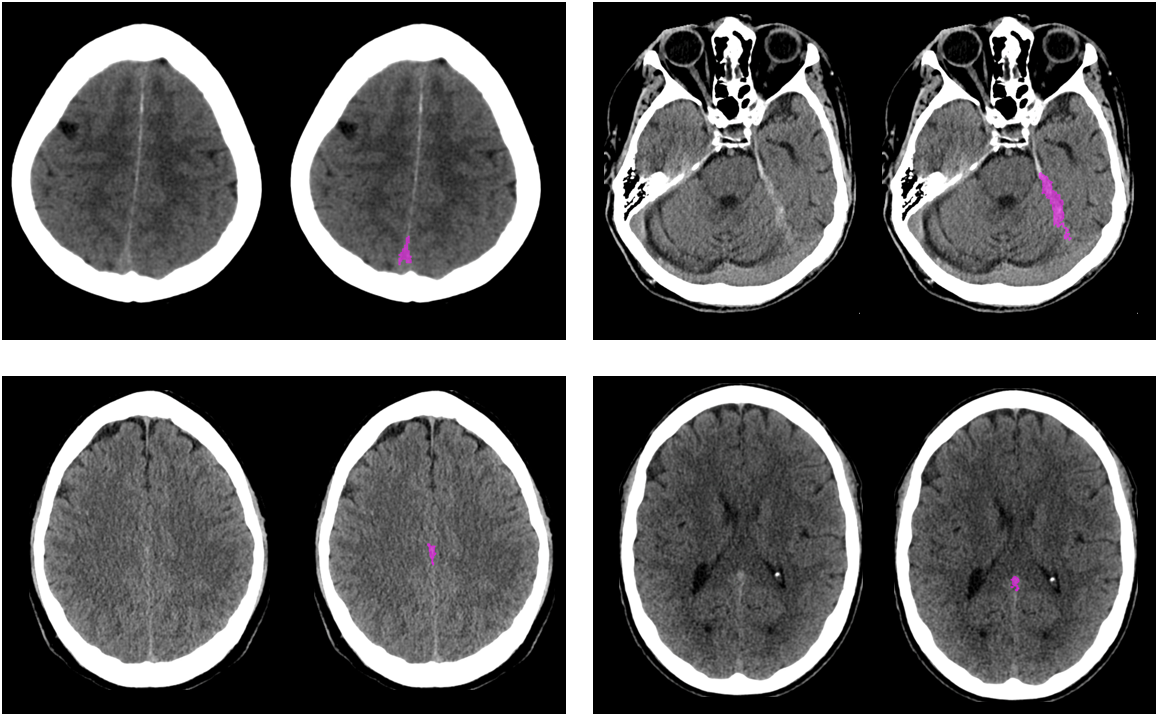

**a)**

**b)**

**c)**

**d)**

**Supplementary Figure 2. Examples of False Positive Segmentations.** Examples of common false positive identifications. **(a)** Superior sagittal sinus, **(b)** Cerebellar tentorium, **(c)** Falx cerebri, and **(d)** Straight sinus. The left-sided image depicts the original image, and the DL solution’s detection is presented in the right-sided image (indicated by the purple color).

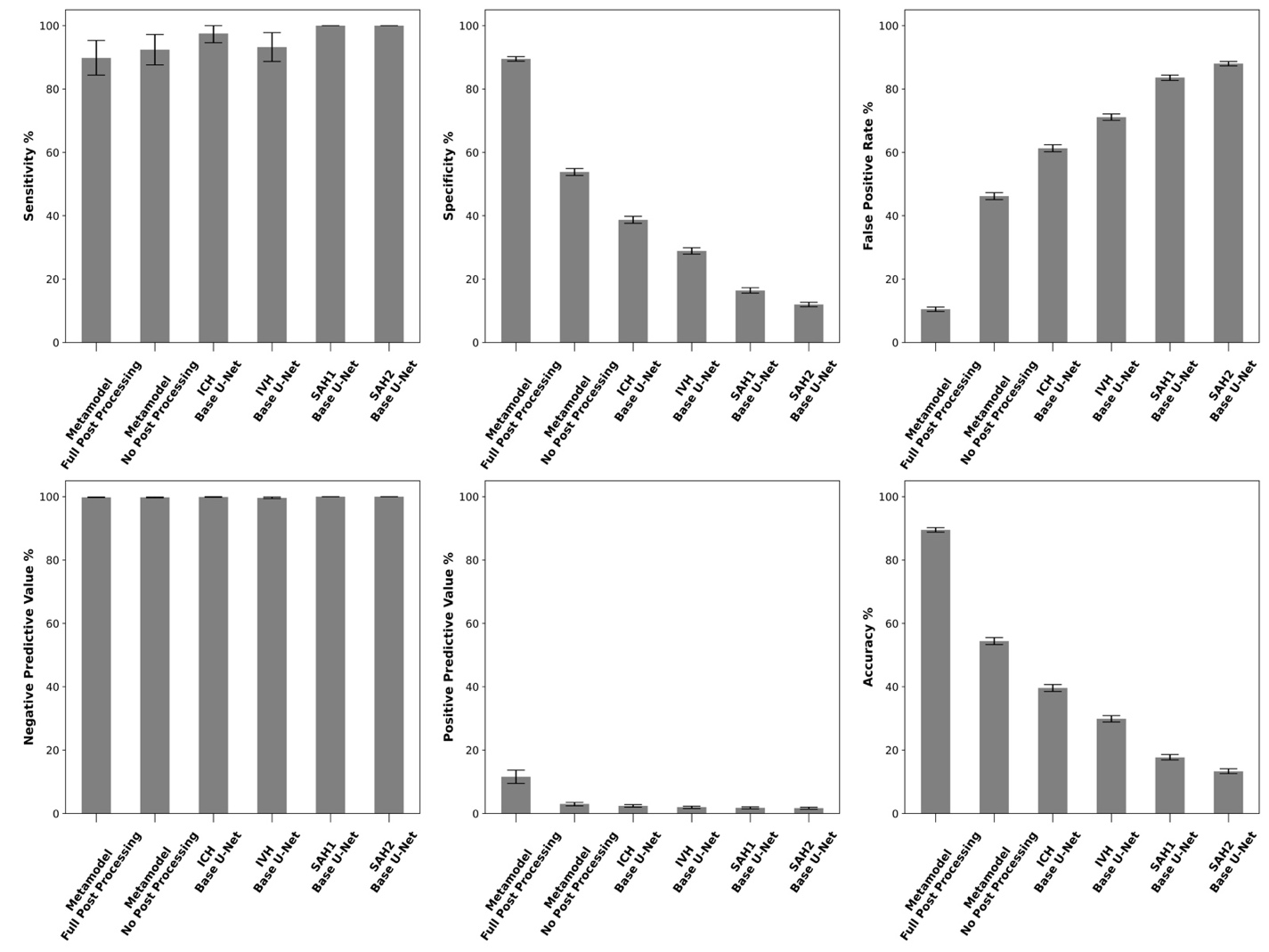

**Supplementary Figure 3. Summary of Case-Level Performances.** Case-level performance of different base U-Nets and metamodel with and without post-processing steps. Bar whiskers represent the 95 % confidence interval for corresponding metric. SAH1 is the new U-Net trained for this study and SAH2 is algorithm described previously by Thanellas et. al.^2^

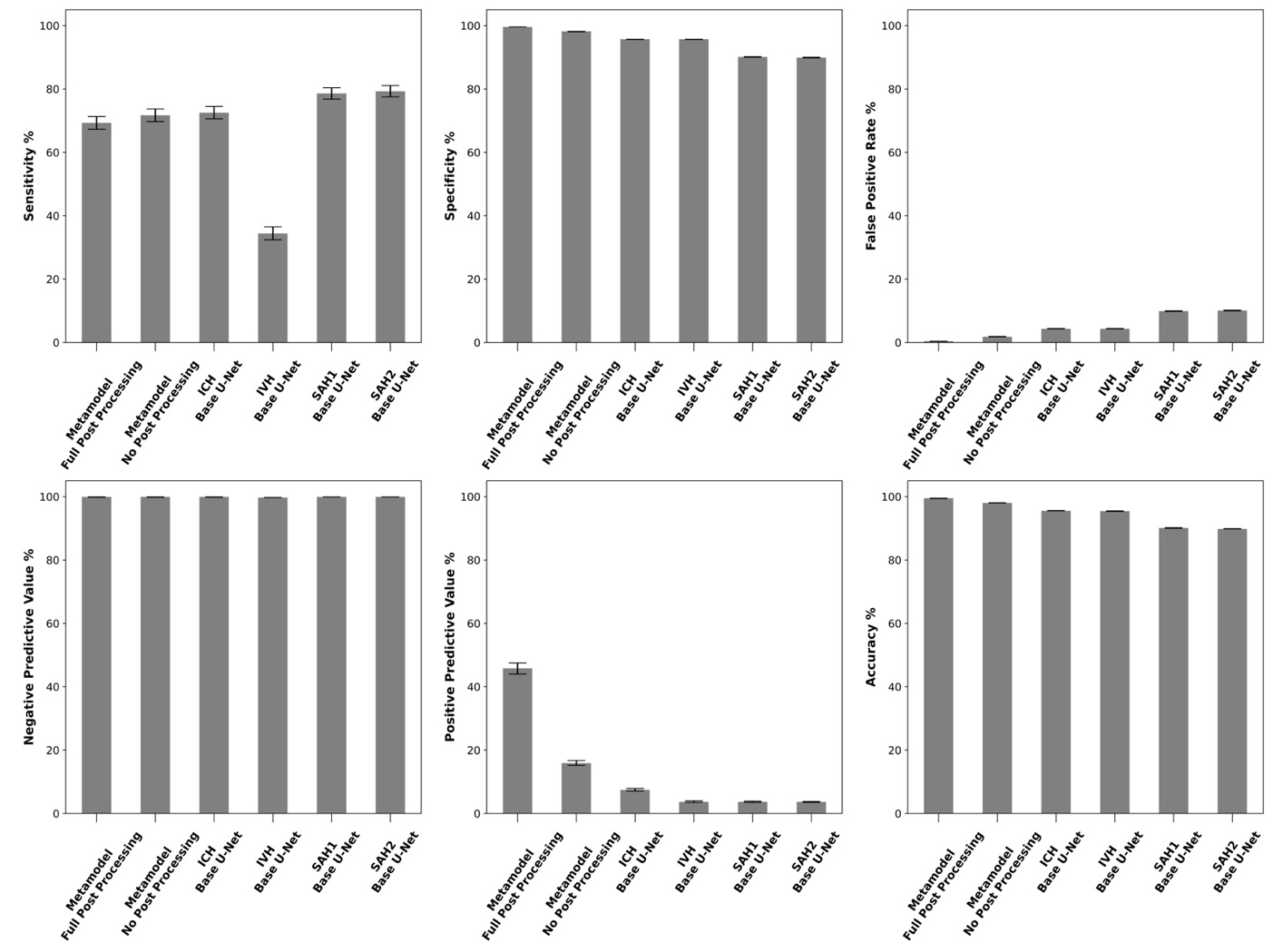

**Supplementary Figure 4. Summary of Slice-Level Performances.** Slice-level performance of different base U-Nets and metamodel with and without post-processing steps. Bar whiskers represent the 95 % confidence interval for corresponding metric. SAH1 is the new U-Net trained for this study and SAH2 is algorithm described previously by Thanellas et. al.^2^

**Missed Hemorrhages**

**Group: Bleed resumed or confirmed to occur within 12–24h**

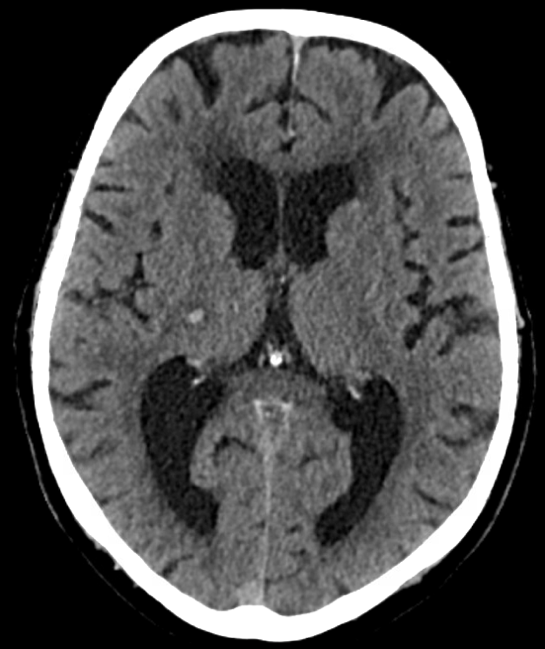

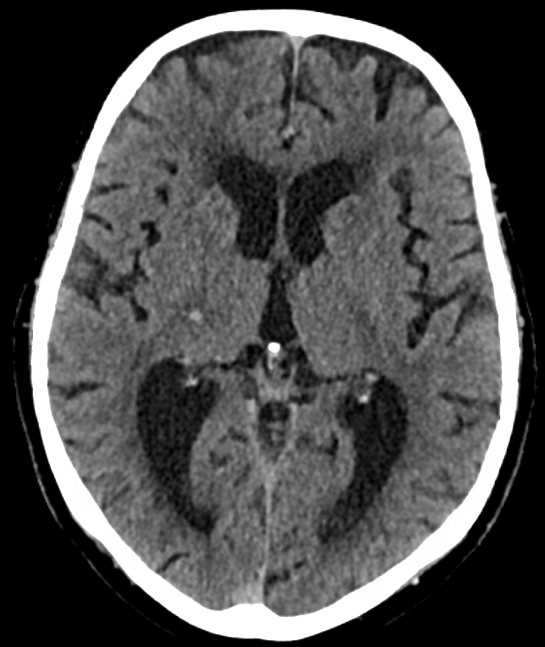

**Supplementary Figure 5.** Small intracerebral hemorrhage in the right internal capsule (indicated by the arrow) with a maximum diameter of 6 mm. The hemorrhage was not detected by our deep learning solutions.

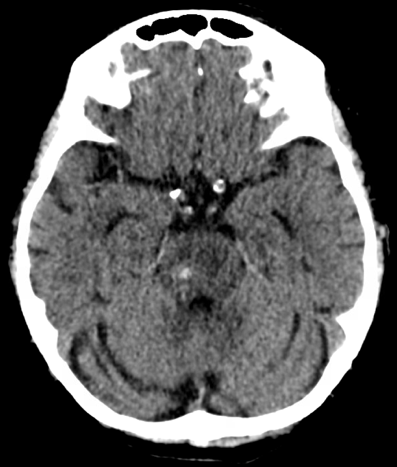

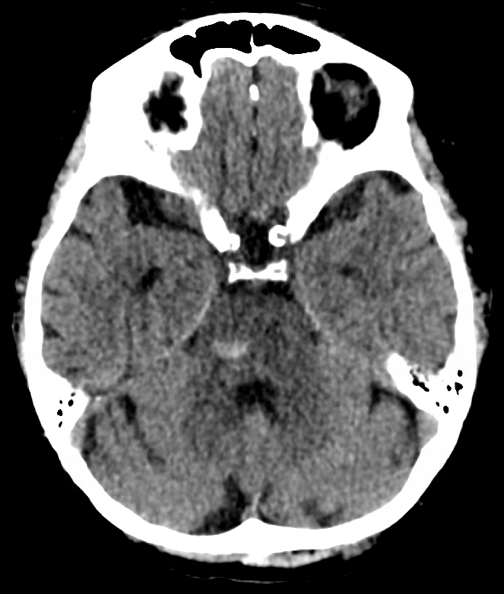

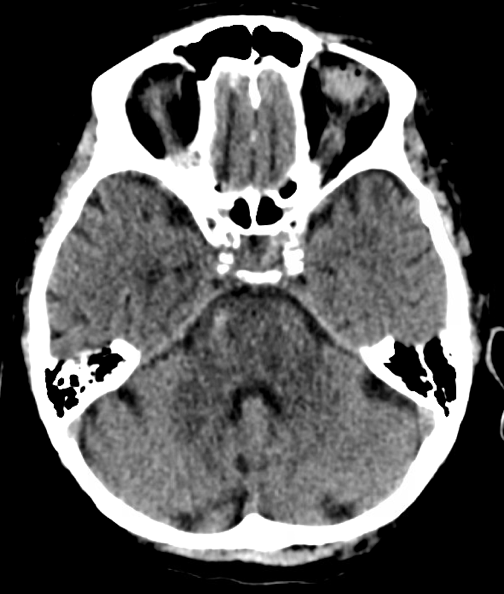

**Supplementary Figure 6.** Intracerebral hemorrhage in the pontine region (indicated by the arrow) with a maximum diameter of 10 mm. The hemorrhage was not detected by our deep learning solution.

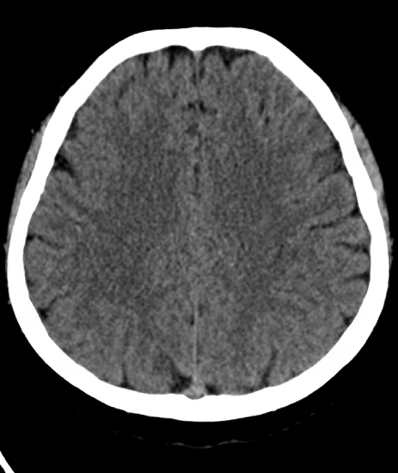

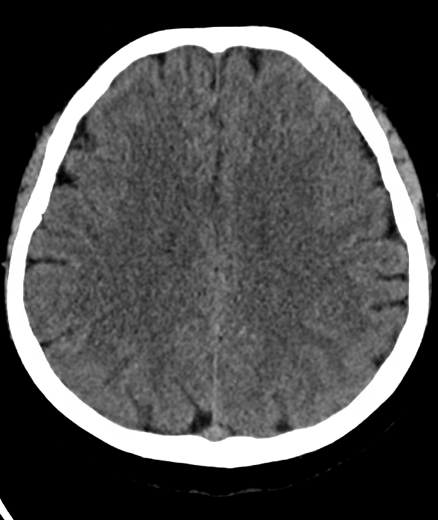

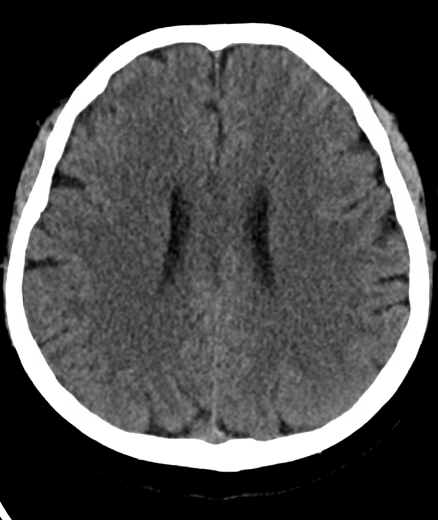

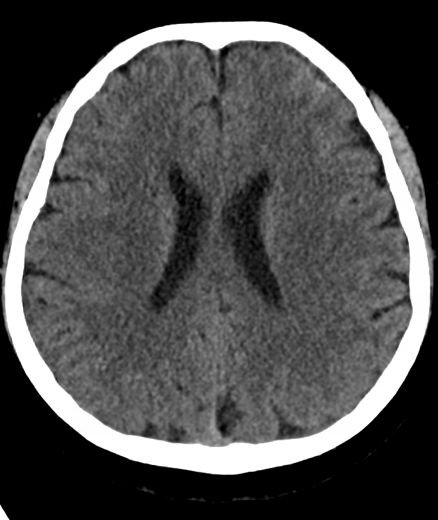

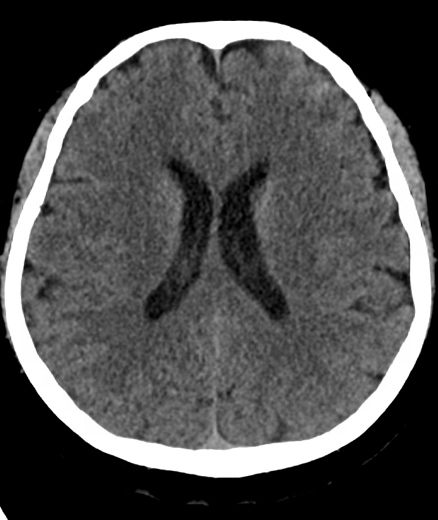

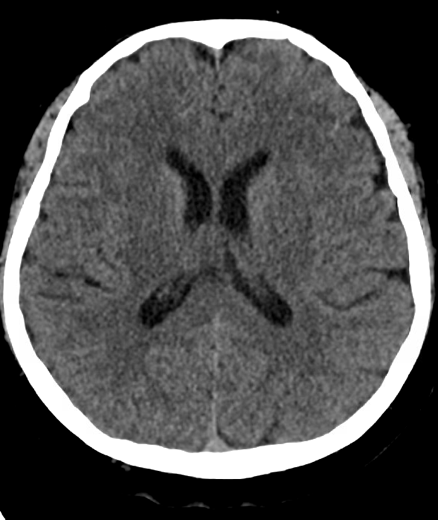

**Supplementary Figure 7.** Left-sided subarachnoid hemorrhage in the sulci (indicated by the arrow). The hemorrhage was not detected by our deep learning solution.

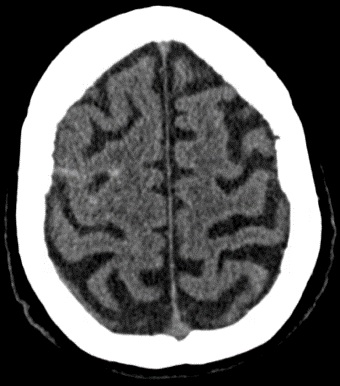

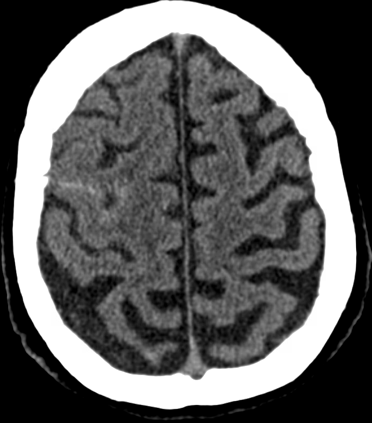

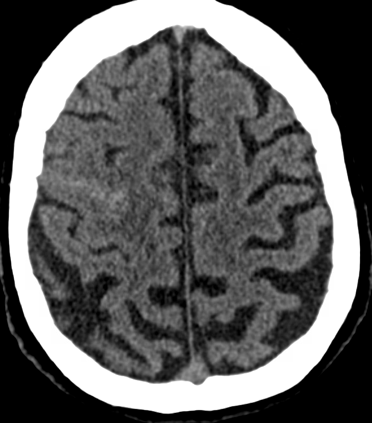

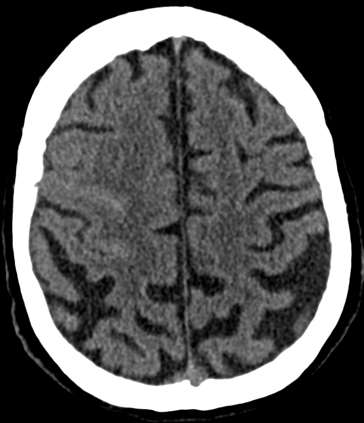

**Supplementary Figure 8.** Right-sided subarachnoid hemorrhage (indicated by the arrow). The hemorrhage was not detected by our deep learning solution.

**Group: Bleed occurred within 24 h–7 days**

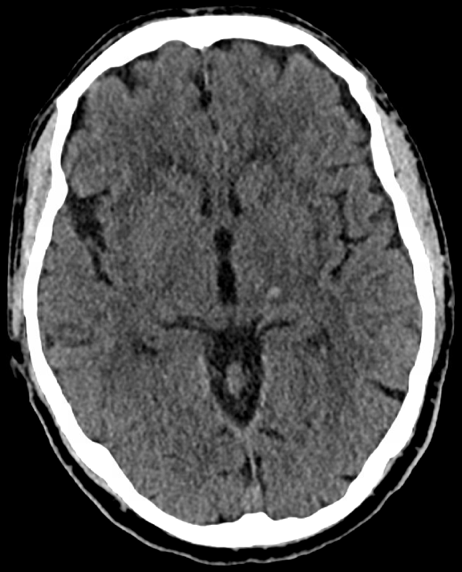

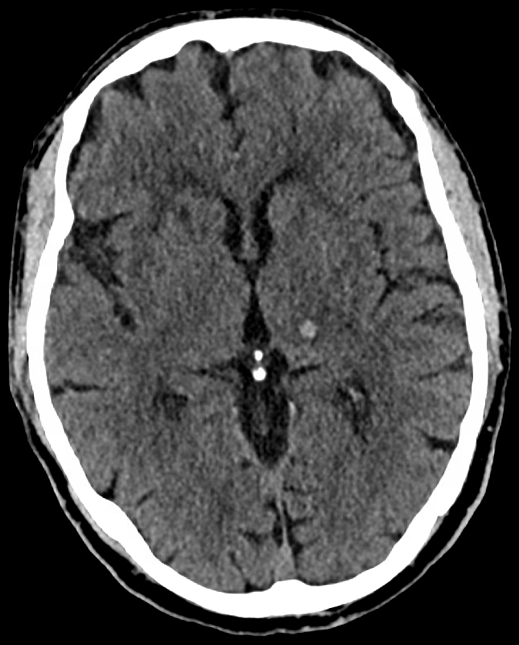

**Supplementary Figure 9.** Small intracerebral hemorrhage in the left internal capsule (indicated by the arrow) with a maximum diameter of 6 mm. The hemorrhage was not detected by our deep learning solution.

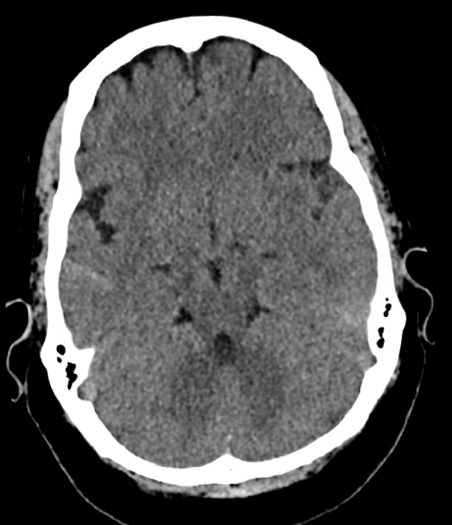

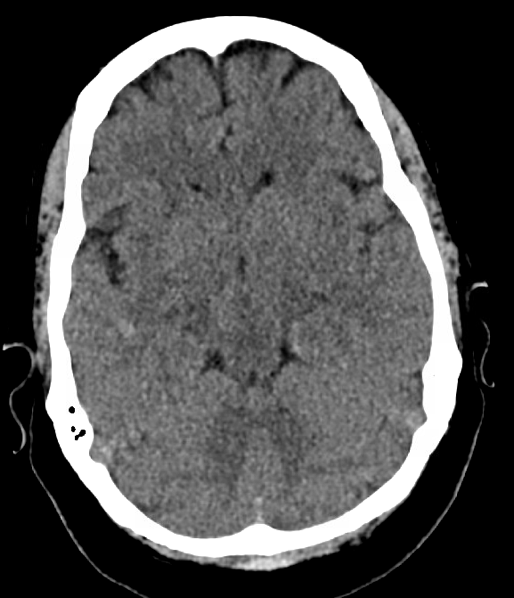

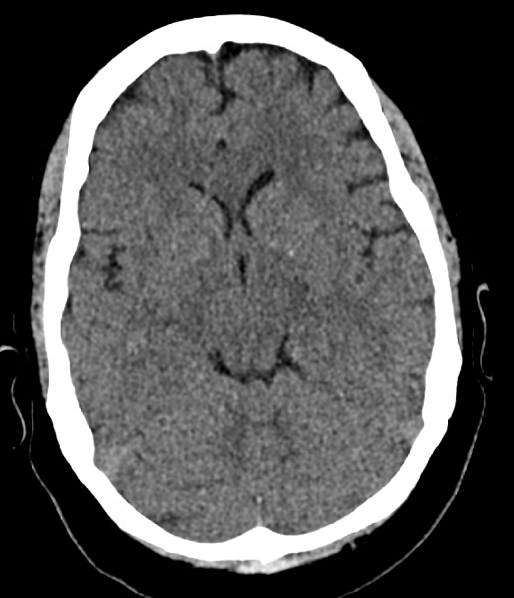

**Supplementary Figure 10.** Right-sided subarachnoid hemorrhage (indicated by the arrow). The hemorrhage was not detected by our deep learning solution.

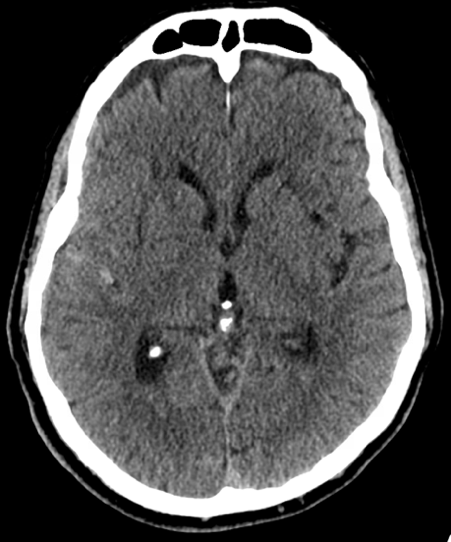

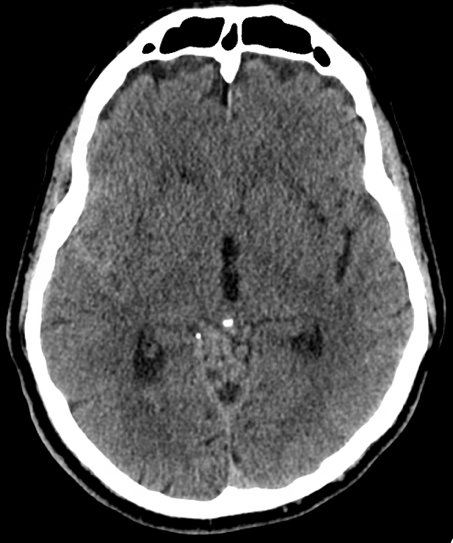

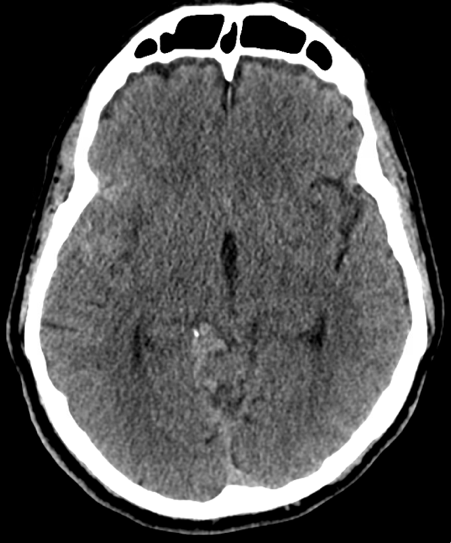

**Supplementary Figure 11.** Right-sided subarachnoid hemorrhage (indicated by the arrow). The hemorrhage was not detected by our deep learning solution.

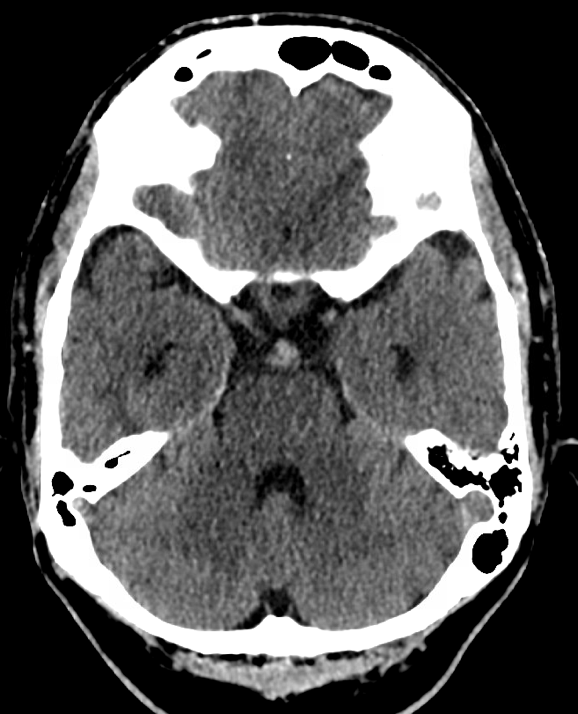

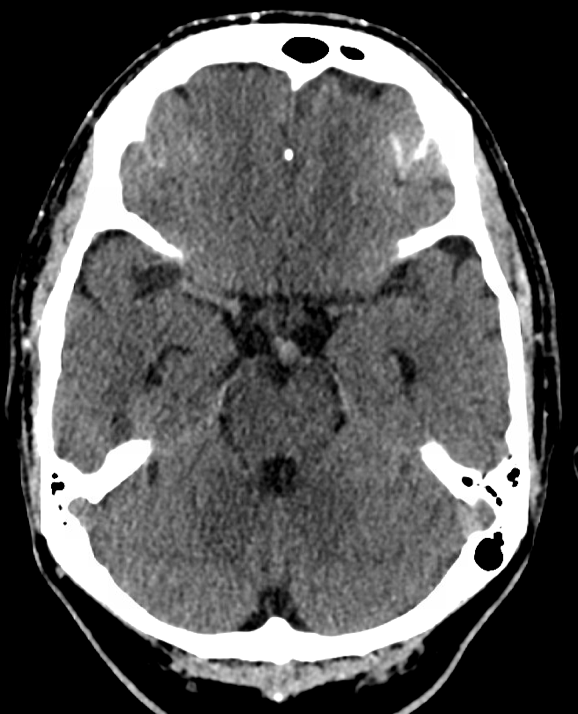

**Supplementary Figure 12.** On-call radiologist reported subarachnoid hemorrhage (indicated by the arrow). The hemorrhage was not detected by our deep learning solution. On-call radiologist report also mentioned that a calcified basilar artery aneurysm would be a less probable differential diagnostic option for the finding.

**Group: Bleed occurred > 14 Days, or Onset Time Unclear**

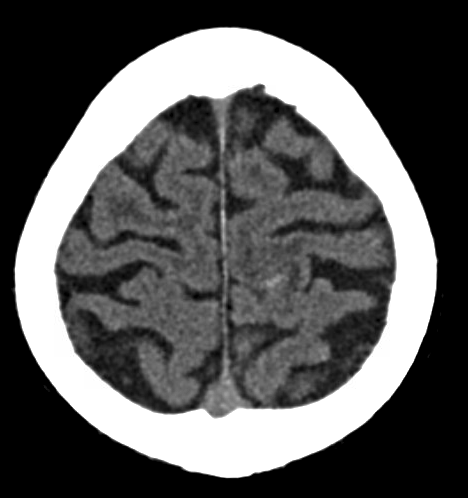

**Supplementary Figure 13.** Small remnant of resorbing intracerebral hemorrhage (indicated by the arrow) with a maximum diameter of 5 mm. The hemorrhage was not detected by our deep learning solution.

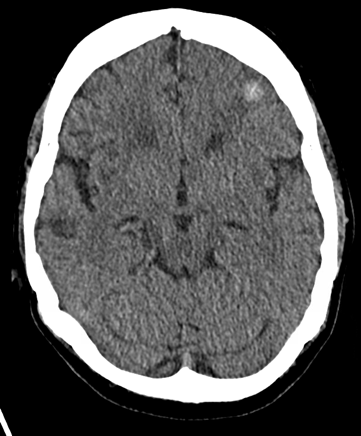

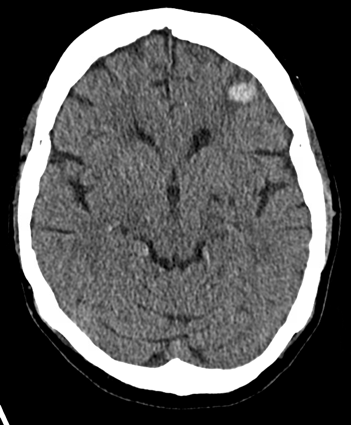

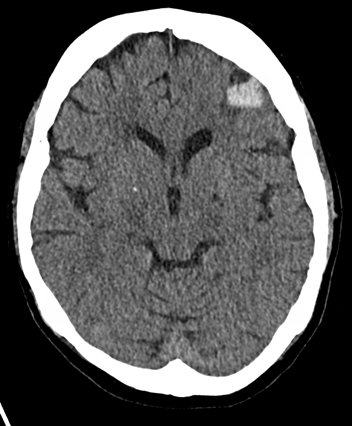

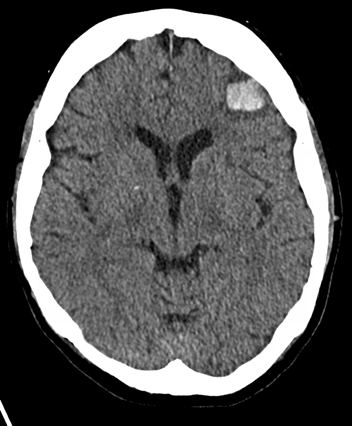

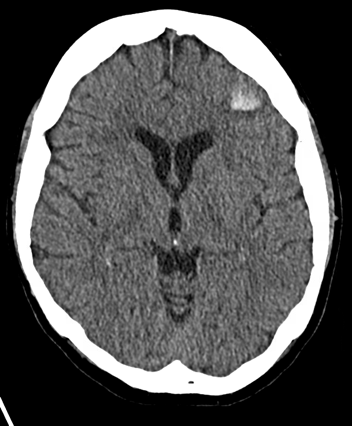

**Supplementary Figure 14.** Left-sided frontal intracerebral hemorrhage with a maximum diameter of 19 mm. The hemorrhage was not detected by our deep learning solution.

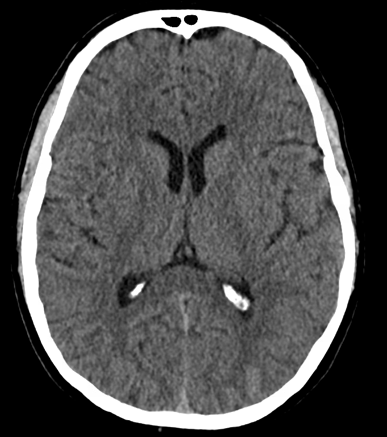

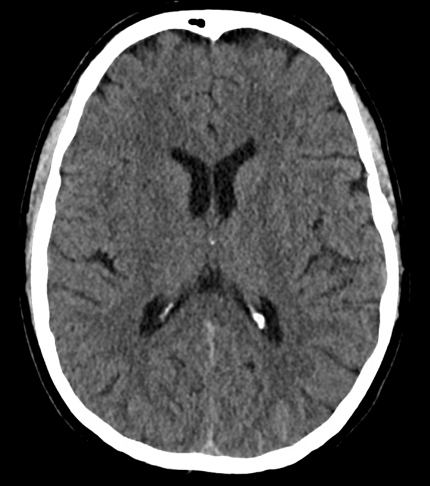

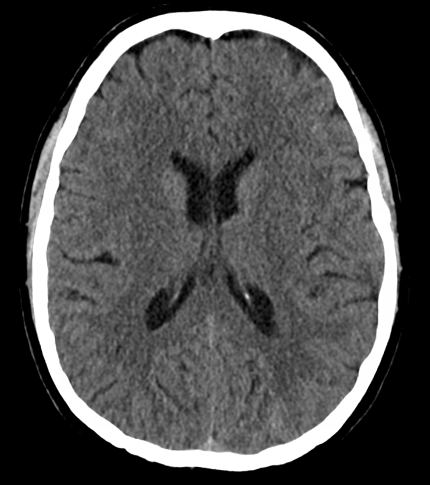

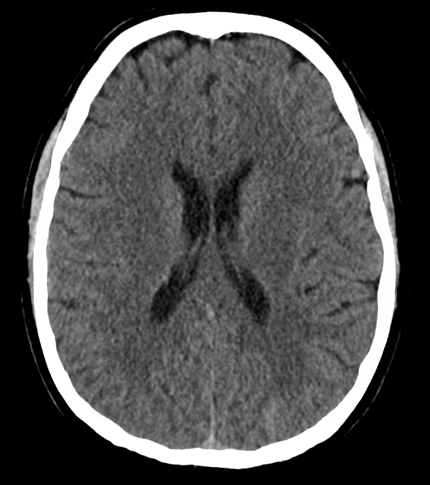

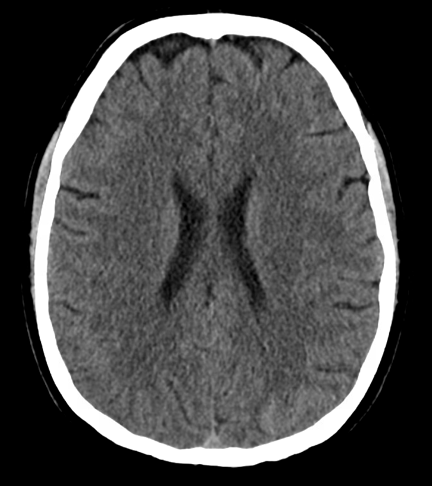

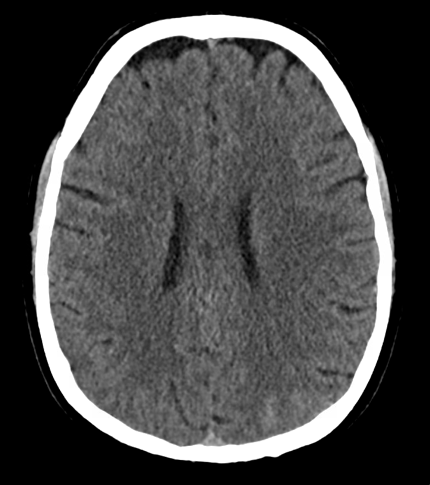

**Supplementary Figure 15.** Left-sided posterior subarachnoid hemorrhage (indicated by the arrow). The hemorrhage was not detected by our deep learning solution.

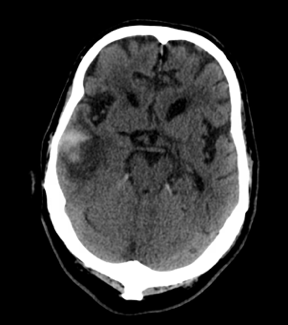

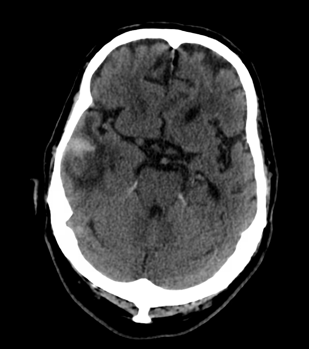

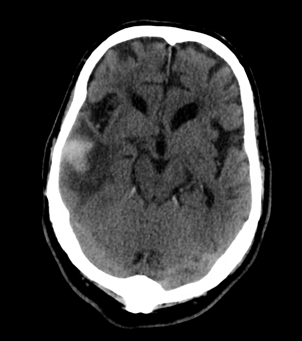

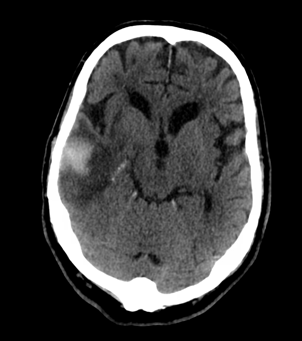

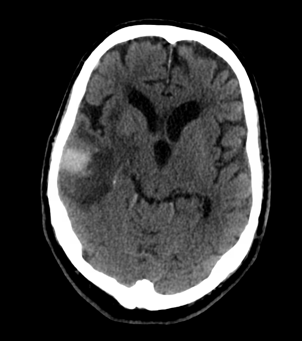

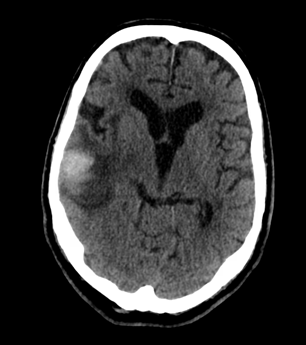

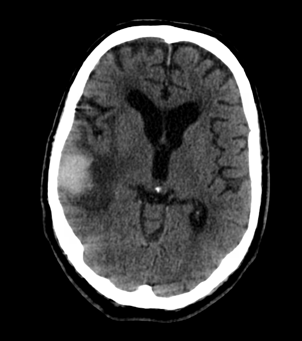

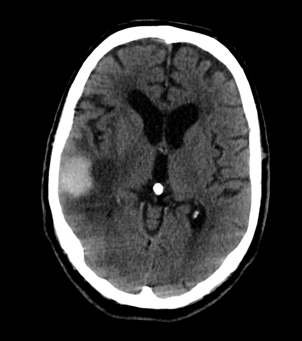

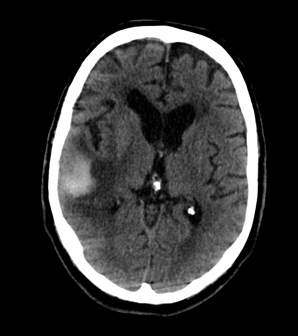

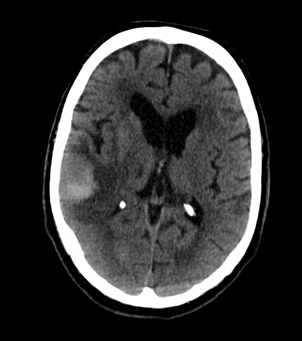

**Supplementary Figure 16.** Right-sided intracerebral hemorrhage in the temporal region with a maximum diameter of 29 mm. The hemorrhage was not detected by our deep learning solution.

**Identified Hemorrhages Missed in On-Call Reports**

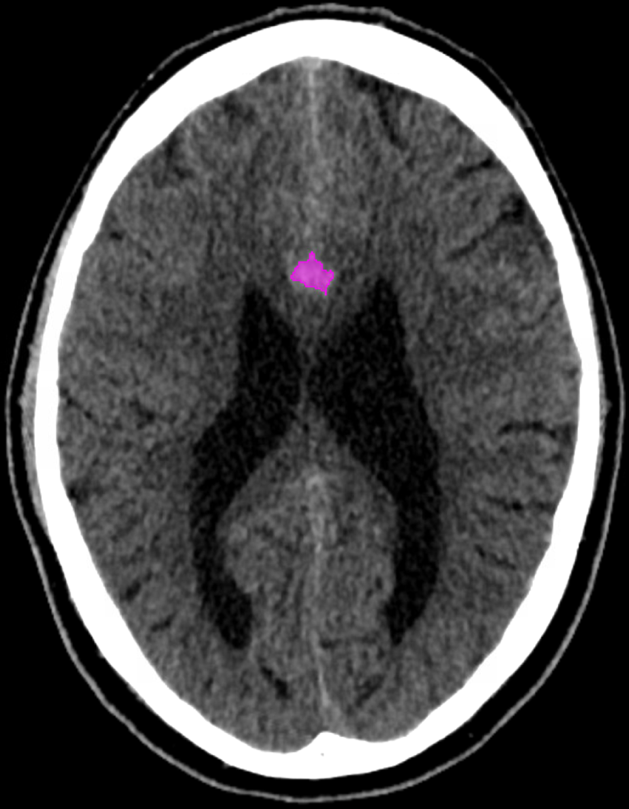

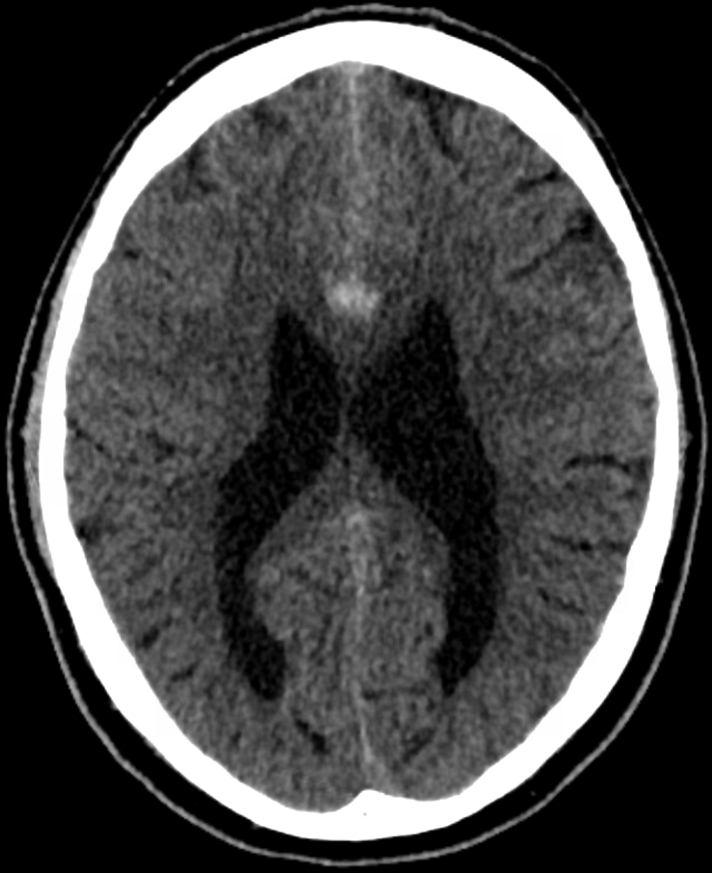

**Original image**

**DL solution’s detection**

**Supplementary Figure 17. Identified Hemorrhage 1.** The NCCT showed a small frontal SAH. The on-call report was negative for hemorrhage. Presence of the SAH was later reported in an additional report by the consultant radiologist. Computed tomography angiography (CTA) was later carried out and showed a pericallosal aneurysm, thus the SAH was considered aneurysmatic.

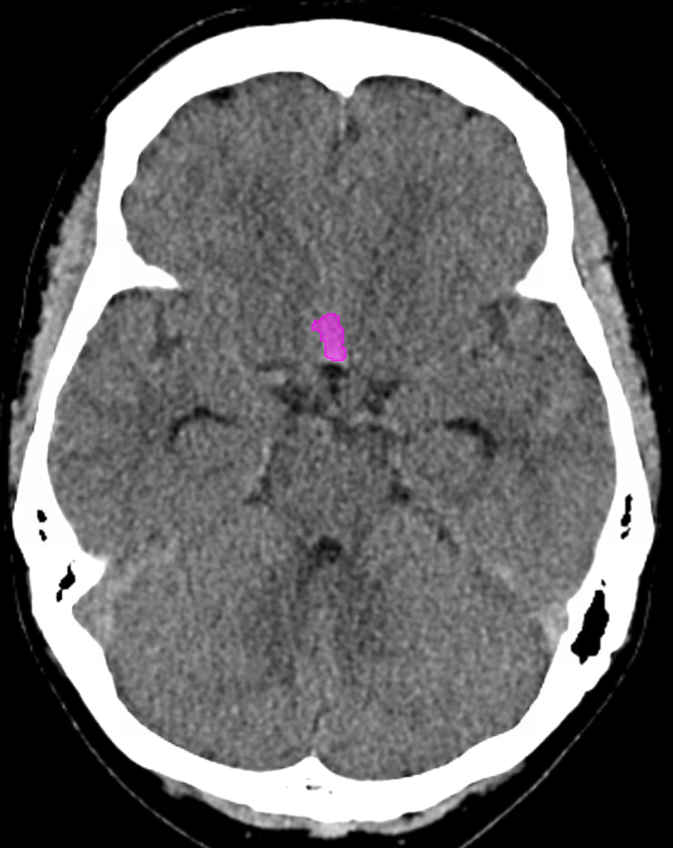

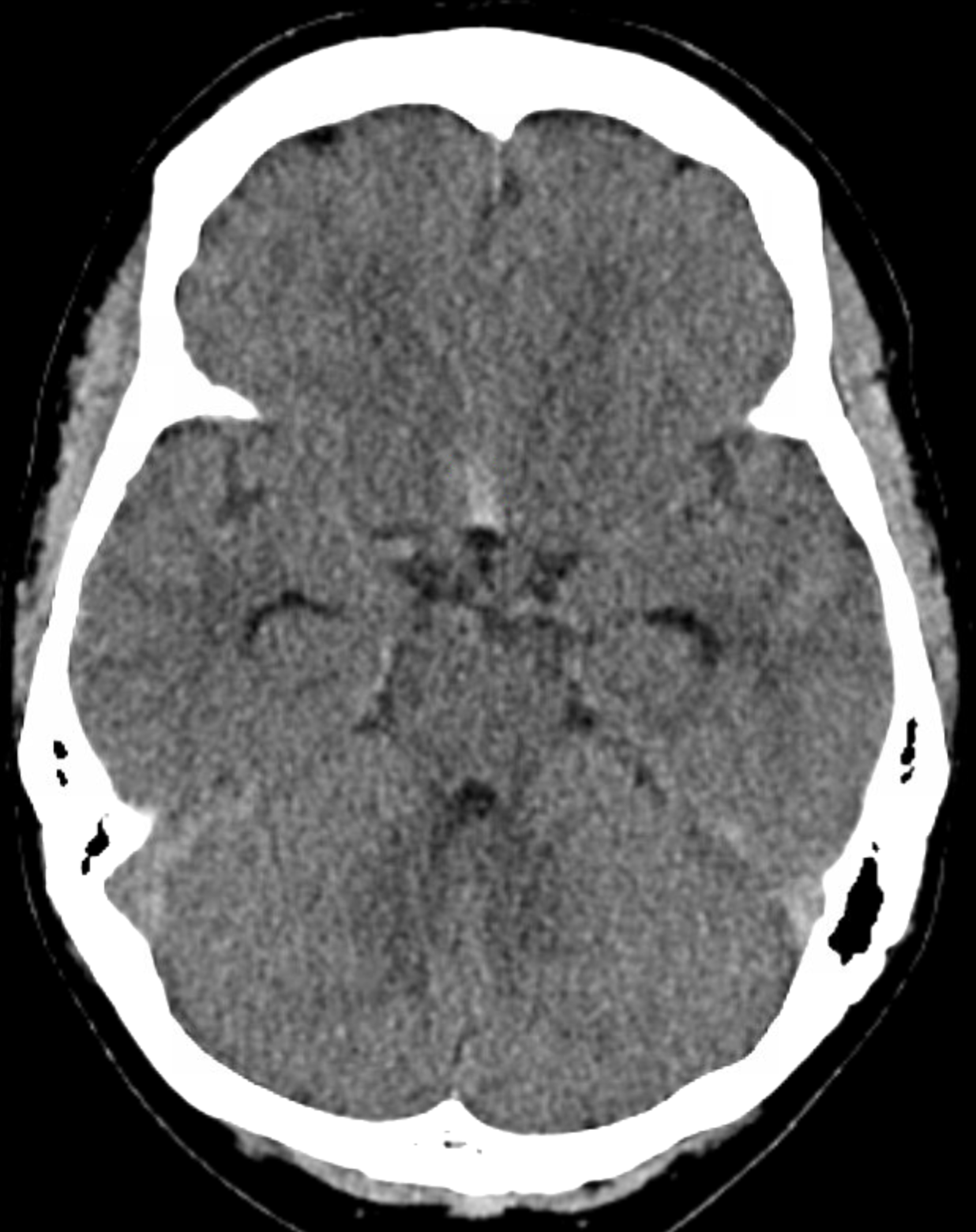

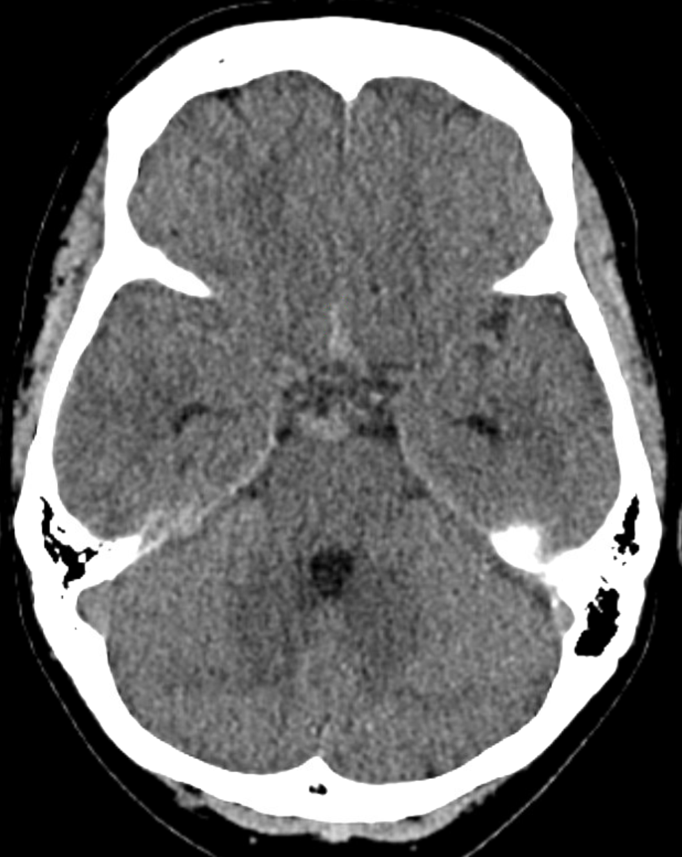

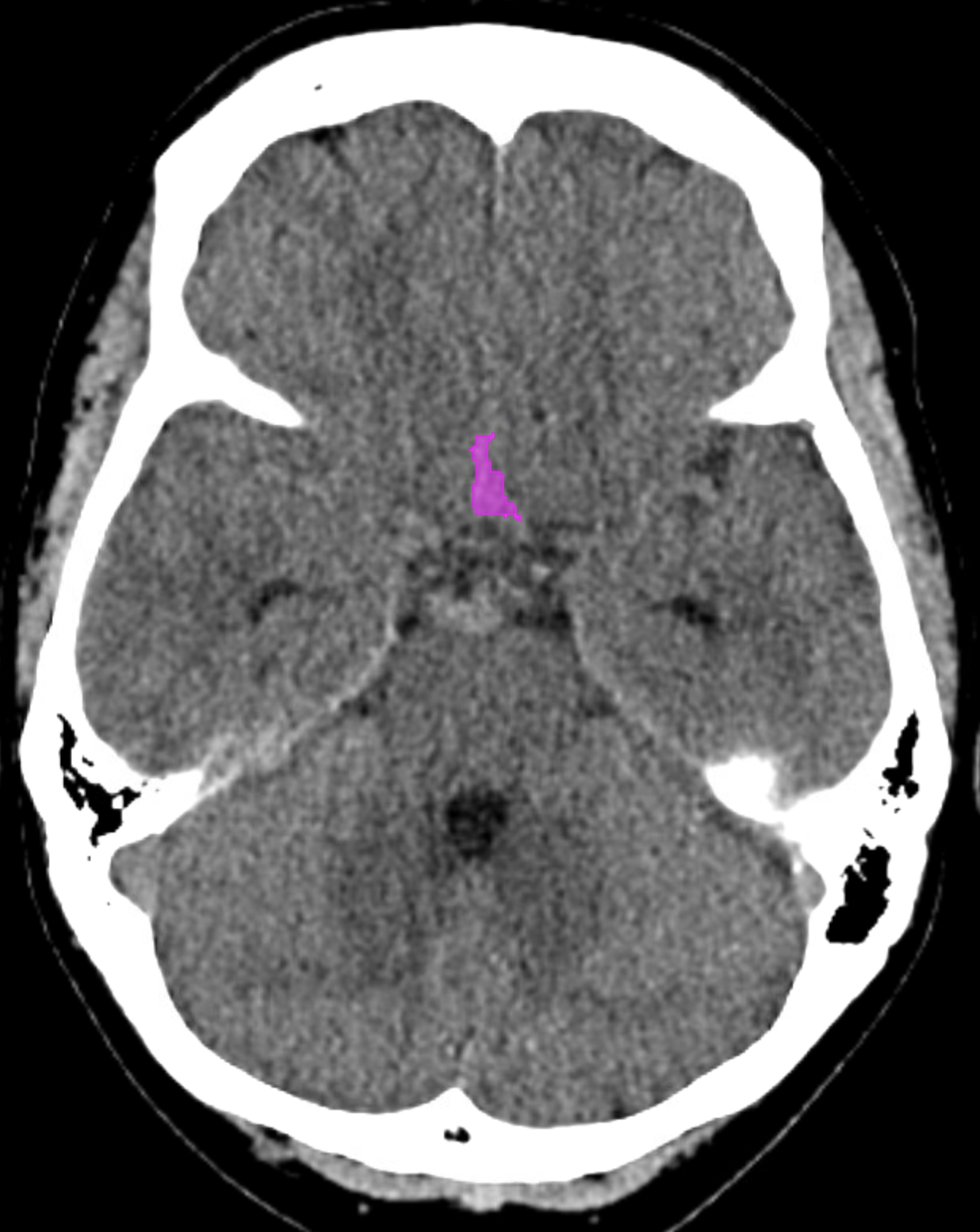

**Original image**

**DL solution’s detection**

**Supplementary Figure 18. Identified Hemorrhage 2.** NCCT showed SAH in the interhemispheric fissure. The initial on-call report was negative for hemorrhage. The SAH was diagnosed with a lumbar puncture. CTA was done as an additional imaging study, and did not reveal an aneurysm, thus the SAH was considered non-aneurysmatic.

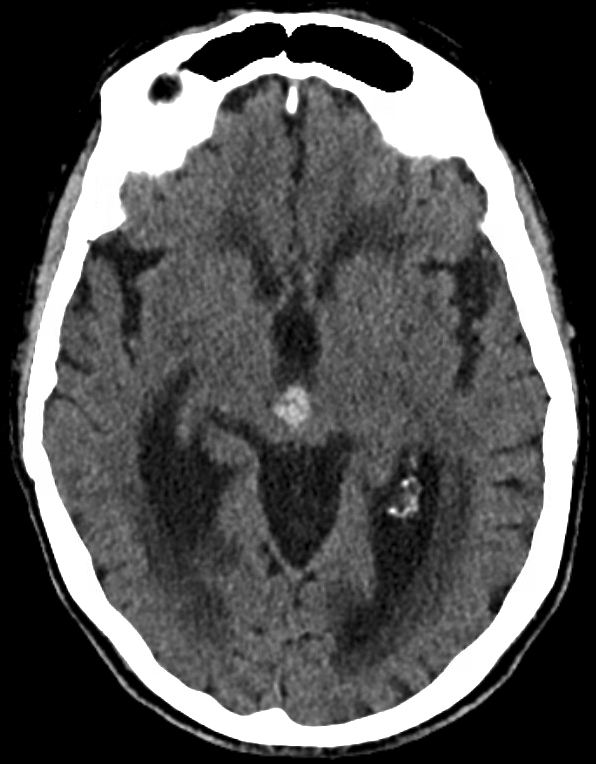

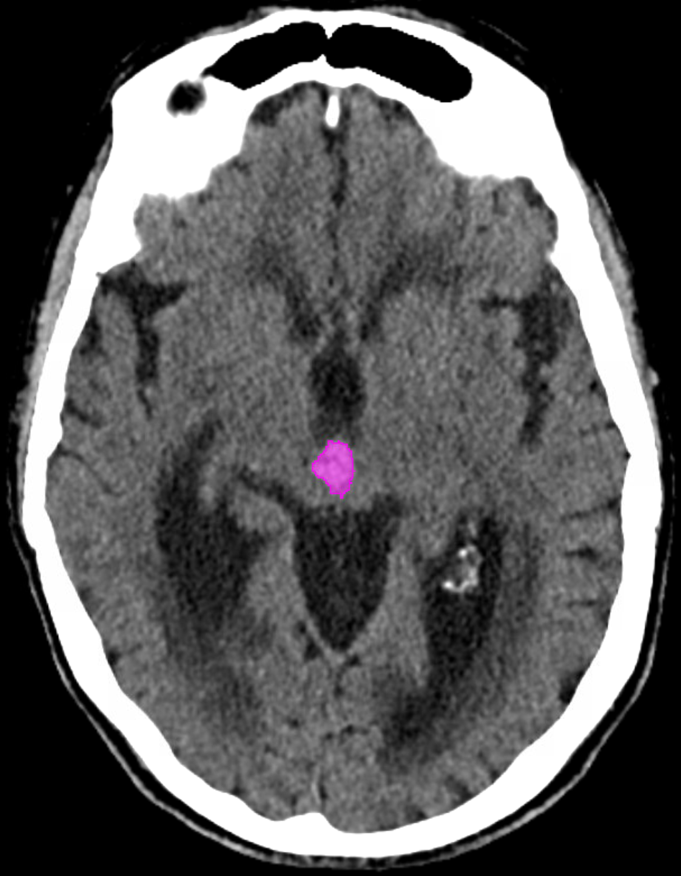

**Original image**

**DL solution’s detection**

**Supplementary Figure 19. Identified Hemorrhage 3.** On-call report stated the NCCT was negative for hemorrhage. Later, an additional report by the consultant radiologist stated an intraventricular hemorrhage and disturbance of cerebrospinal fluid circulation.

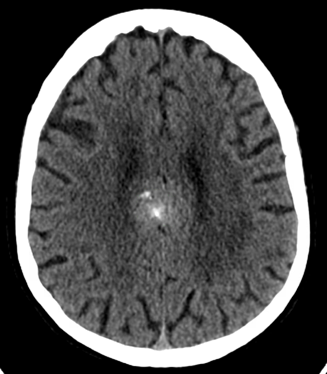

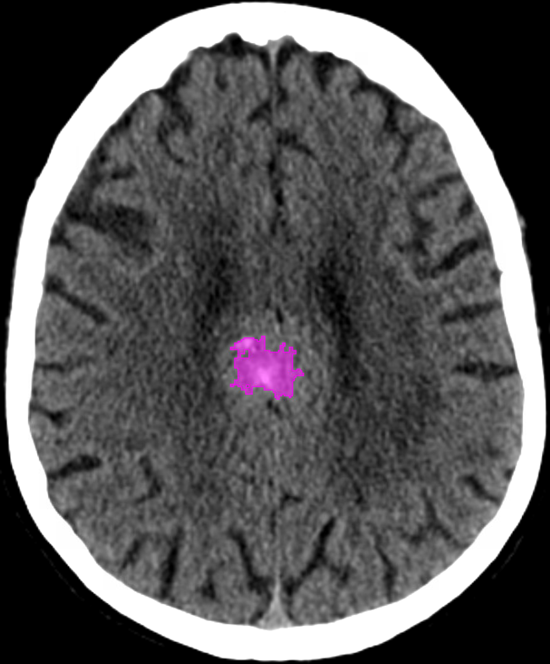

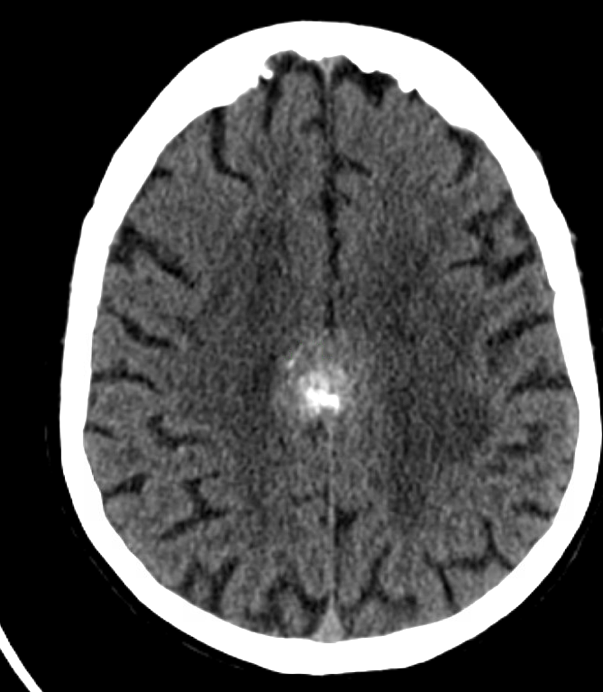

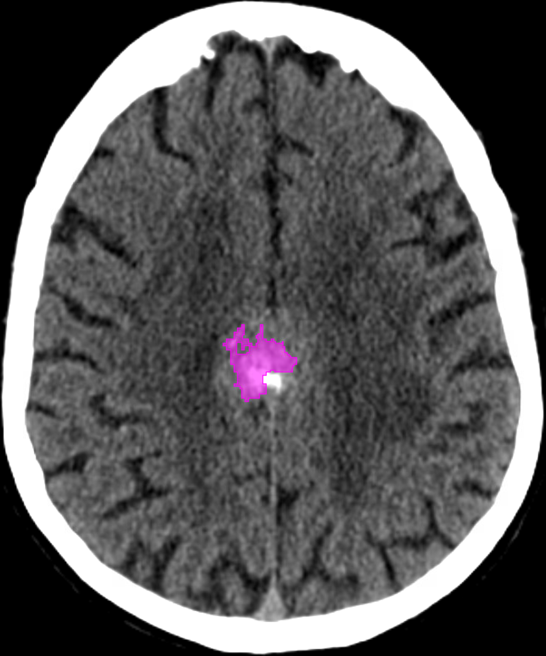

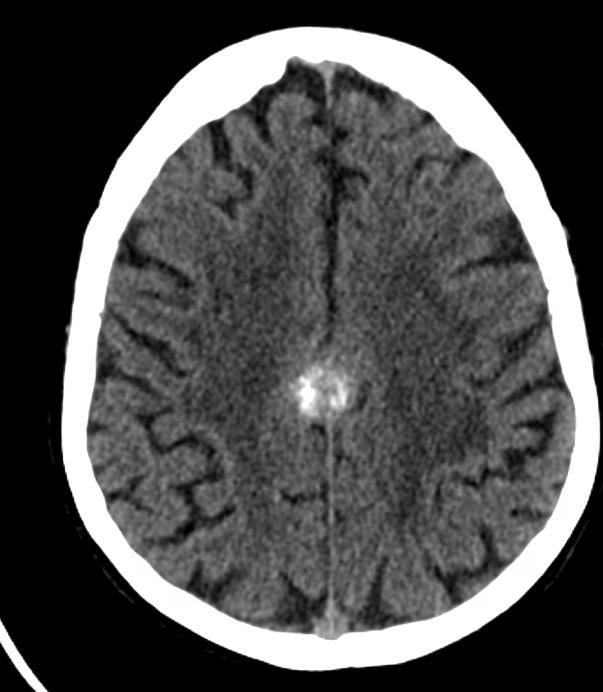

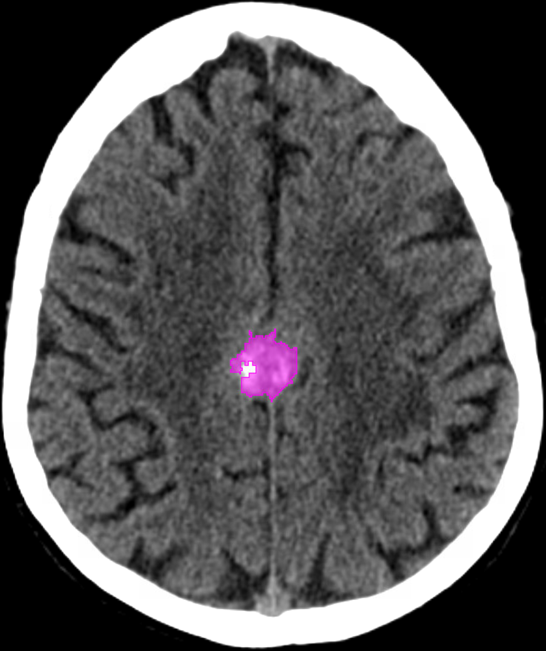

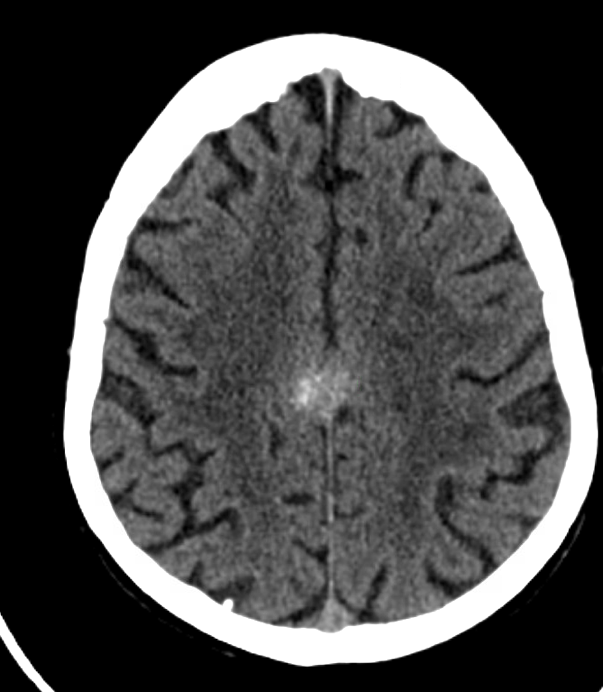

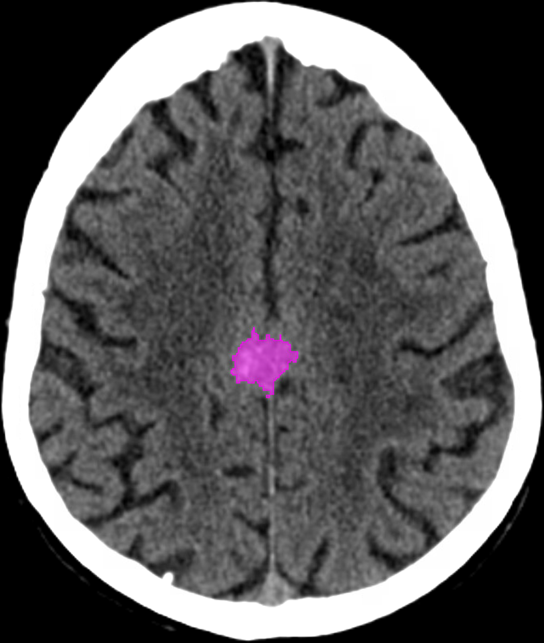

**Original image**

**DL solution’s detection**

**Original image**

**DL solution’s detection**

**Supplementary Figure 20. Identified Hemorrhage 4.** On-call radiologist initially reported the finding as meningioma, but later revised the report and stated that the finding included hyperdense regions, suggesting that the finding was intracranial bleeding instead of meningioma.

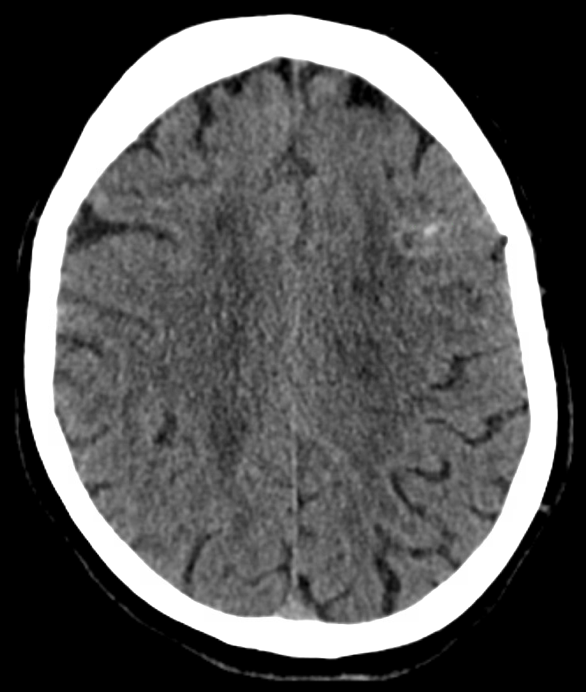

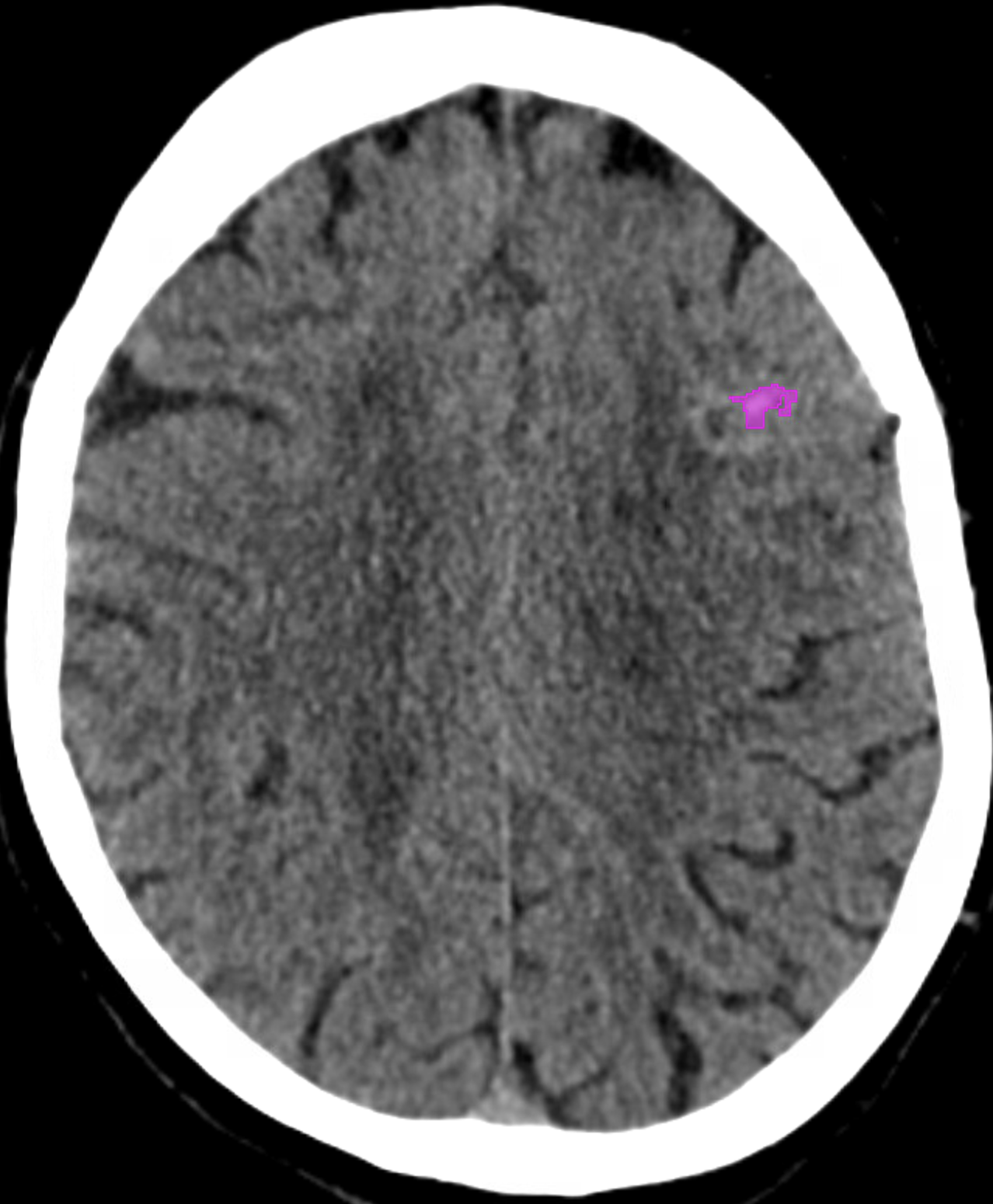

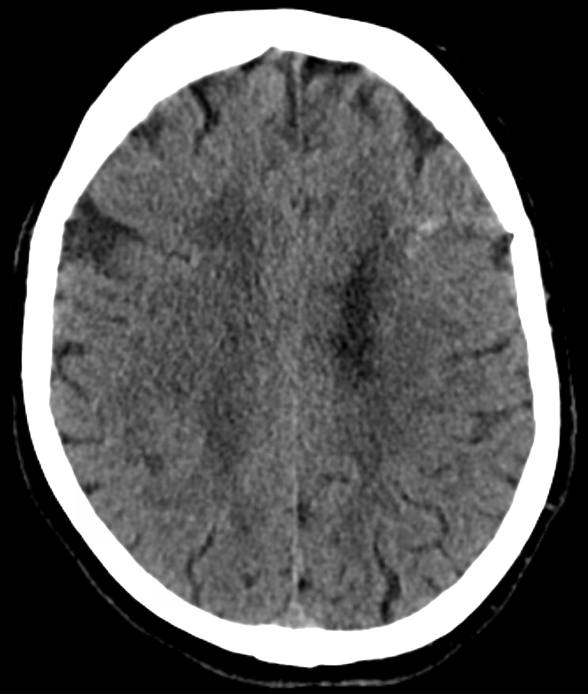

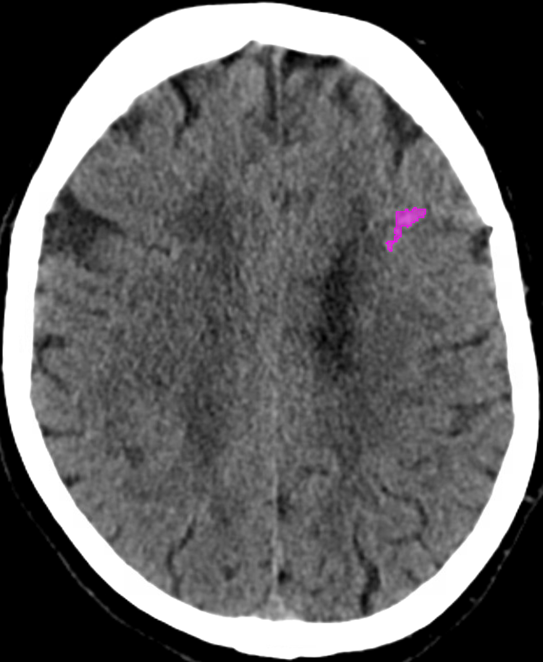

**Original image**

**DL solution’s detection**

**Supplementary Figure 21. Identified Hemorrhage 5**. On-call report stated the finding as calcification related to resorbing subacute hemorrhage. Later, an additional radiologist report by the consultant radiologist stated that the finding was a hemorrhage
